# JAK inhibition decreases the autoimmune burden in Down syndrome

**DOI:** 10.1101/2024.06.13.24308783

**Authors:** Angela L. Rachubinski, Elizabeth Wallace, Emily Gurnee, Belinda A. Enriquez Estrada, Kayleigh R. Worek, Keith P. Smith, Paula Araya, Katherine A. Waugh, Ross E. Granrath, Eleanor Britton, Hannah R. Lyford, Micah G. Donovan, Neetha Paul Eduthan, Amanda A. Hill, Barry Martin, Kelly D. Sullivan, Lina Patel, Deborah J. Fidler, Matthew D. Galbraith, Cory A. Dunnick, David A. Norris, Joaquin M. Espinosa

**Author notes:** Corresponding authors /.

## Abstract

Individuals with Down syndrome (DS), the genetic condition caused by trisomy 21 (T21), display clear signs of immune dysregulation, including high rates of autoimmune disorders and severe complications from infections. Although it is well established that T21 causes increased interferon responses and JAK/STAT signaling, elevated autoantibodies, global immune remodeling, and hypercytokinemia, the interplay between these processes, the clinical manifestations of DS, and potential therapeutic interventions remain ill defined. Here, we report a comprehensive analysis of immune dysregulation at the clinical, cellular, and molecular level in hundreds of individuals with DS. We demonstrate multi-organ autoimmunity of pediatric onset concurrent with unexpected autoantibody-phenotype associations. Importantly, constitutive immune remodeling and hypercytokinemia occur from an early age prior to autoimmune diagnoses or autoantibody production. We then report the interim analysis of a Phase II clinical trial investigating the safety and efficacy of the JAK inhibitor tofacitinib through multiple clinical and molecular endpoints. Analysis of the first 10 participants to complete the 16-week study shows a good safety profile and no serious adverse events. Treatment reduced skin pathology in alopecia areata, psoriasis, and atopic dermatitis, while decreasing interferon scores, cytokine scores, and levels of pathogenic autoantibodies without overt immune suppression. Additional research is needed to define the effects of JAK inhibition on the broader developmental and clinical hallmarks of DS. ClinicalTrials.gov identifier: NCT04246372.

## Introduction

Trisomy of human chromosome 21 (T21) occurs at a rate of ∼1 in 700 live births, causing Down syndrome (DS)^1,2^. Individuals with DS display a distinct clinical profile including developmental delays, stunted growth, cognitive impairments, and increased risk of leukemia, autism spectrum disorders, seizure disorders, and Alzheimer’s disease^2,3^. People with DS also display widespread immune dysregulation, which manifests through severe complications from respiratory viral infections and high prevalence of myriad immune conditions, including autoimmune thyroid disease (AITD)^4–6^, celiac disease^7,8^, and skin conditions such as atopic dermatitis, alopecia areata, hidradenitis suppurativa (HS), vitiligo, and psoriasis^9–11^. Furthermore, people with DS display signs of neuroinflammation from an early age^12–14^. Although it is now well accepted that immune dysregulation is a hallmark of DS, the underlying mechanisms and therapeutic implications are not yet fully defined.

We previously reported that T21 causes consistent activation of the interferon (IFN) transcriptional response in multiple immune and non-immune cell types with concurrent hypersensitivity to IFN stimulation and hyperactivation of downstream JAK/STAT signaling^15–17^. Plasma proteomics studies identified dozens of inflammatory cytokines with mechanistic links to IFN signaling that are elevated in people with DS^18^. A large metabolomics study revealed that T21 drives the production of neurotoxic tryptophan catabolites via the IFN-inducible kynurenine pathway^19^. Deep immune profiling revealed global immune remodeling with hypersensitivity to IFN across all major branches of the immune system^15^, and dysregulation of T cell lineages toward a hyperactive, autoimmunity-prone state^16^. These results could be partly explained by the fact that four of the six IFN receptors (IFNRs) are encoded on chr21, including Type I, II and III IFNR subunits^20^. In a mouse model of DS, normalization of *IFNR* gene copy number rescues multiple phenotypes of DS, including lethal immune hypersensitivity, congenital heart defects (CHDs), cognitive impairments, and craniofacial anomalies^21^. JAK inhibition rescues lethal immune hypersensitivity in these mouse models^22^ and attenuates the global dysregulation of gene expression caused by the trisomy across multiple murine tissues^23^. Furthermore, prenatal JAK inhibition in pregnant mice prevents the appearance of CHDs^24^. Altogether, these results support the notion that T21 elicits an interferonopathy in DS, and that pharmacological inhibition of IFN signaling could have multiple therapeutic benefits in this population.

Although it is now well established that T21 disrupts immune homeostasis toward an autoimmunity-prone state, the interplay between overexpression of chromosome 21 genes, hyperactive interferon signaling, dysregulation of immune cell lineages, autoantibody production, hypercytokinemia, and the various developmental and clinical features of DS remain to be elucidated. Previous studies established similarities between the immune profiles of typical aging, autoimmunity in the general population, and DS, proposing a role for accelerated immune aging in the pathophysiology of DS^25–27^. Other studies indicate a role for elevated cytokine production, hyperactivated T cells, and ongoing B cell activation as drivers of autoimmunity in DS^15,16,28^. However, given the relatively small sample sizes and observational nature of these studies, it has not been possible to define the contribution of specific dysregulated events to breach of tolerance leading to clinically evident autoimmunity in DS. Therefore, additional research is needed to define driver versus bystander events that could illuminate therapeutic strategies to decrease the burden of autoimmunity in DS.

Within this context, we report here a comprehensive analysis of the immune disorder of DS, including detailed annotation of autoimmune and inflammatory conditions and quantification of autoantibodies in hundreds of research participants, which reveals widespread autoimmune attack on all major organ systems in DS from an early age, including unexpected autoantibody-phenotype associations. Then, using deep immune mapping and quantitative proteomics, we demonstrate that T21 causes widespread immune remodeling toward an autoimmunity-prone state accompanied by hypercytokinemia prior to clinically evident autoimmunity or autoantibody production. Lastly, we report the interim analysis of a clinical trial investigating the safety and efficacy of the JAK1/3 inhibitor tofacitinib (Xeljanz, Pfizer) in DS. These results demonstrate that JAK inhibition improves multiple immunodermatological conditions in DS, normalizes interferon scores, decreases levels of major pathogenic cytokines (e.g., TNF-α, IL-6), and reduces levels of pathogenic autoantibodies [e.g., anti-thyroid peroxidase (anti-TPO)]. Altogether, these results point to hyperactive JAK/STAT signaling as driver of autoimmunity in DS and justify the ongoing trials of JAK inhibitors in DS for multiple clinical endpoints.

## Results

### Widespread multi-organ autoimmunity and autoantibody production in Down syndrome

Previous studies have documented increased rates of diverse autoimmune conditions in DS relative to the general population including autoimmune thyroid disease (AITD)^29^, celiac disease^30^, autoimmune skin conditions^9–11^, and type I diabetes^31,32^. However, many of these studies were limited by relatively small sample sizes, independent analysis of individual autoimmune conditions, or a focus on specific age ranges. In order to complete a more comprehensive analysis of autoimmune conditions in DS across the lifespan, we analyzed the harmonized clinical profiles of 441 research participants with DS, aged 6 months to 57 years, enrolled in the Human Trisome Project cohort study (HTP, NCT02864108), which annotates clinical data through a combination of participant/caregiver surveys and expert abstraction of electronic health records (EHRs) (see **Materials and Methods**, **Supplementary file 1**). In this analysis, the most common autoimmune condition is AITD, affecting 53.1% of the total cohort (**Figure 1a**, **Figure 1 – figure supplement 1a**). Grouped together, autoimmune and inflammatory skin conditions represent the second most common category, affecting 43% of the cohort, including: atopic dermatitis / eczema (27.9%), hidradenitis suppurativa / folliculitis / boils (20.6%), alopecia areata (7.7%), psoriasis (6.1%), and vitiligo (1.9%) (**Figure 1a**, **Figure 1 - supplement 1b**). These observations align with recent epidemiological studies demonstrating high rates of autoimmune and inflammatory skin conditions in DS^33,34^. The rate of celiac disease (9.6%) is also highly elevated over that of the general population^35^. We observed 10 cases (2.2%) of juvenile Type I diabetes, which has been reported to be more common in DS^31,32^. Other autoimmune conditions common in the general population, such as systemic lupus erythematosus or multiple sclerosis, were not observed in the HTP cohort. Other salient conditions annotated in this cohort include recurrent otitis media (15.5%), frequent/recurrent pneumonia (9.2%), severe congenital heart defects requiring surgical repair (19.5%), acute lymphocytic leukemia (ALL, 1.12%), and acute myeloid leukemia (AML, 1.3%) (**Supplementary file 1**).

**Figure 1.**
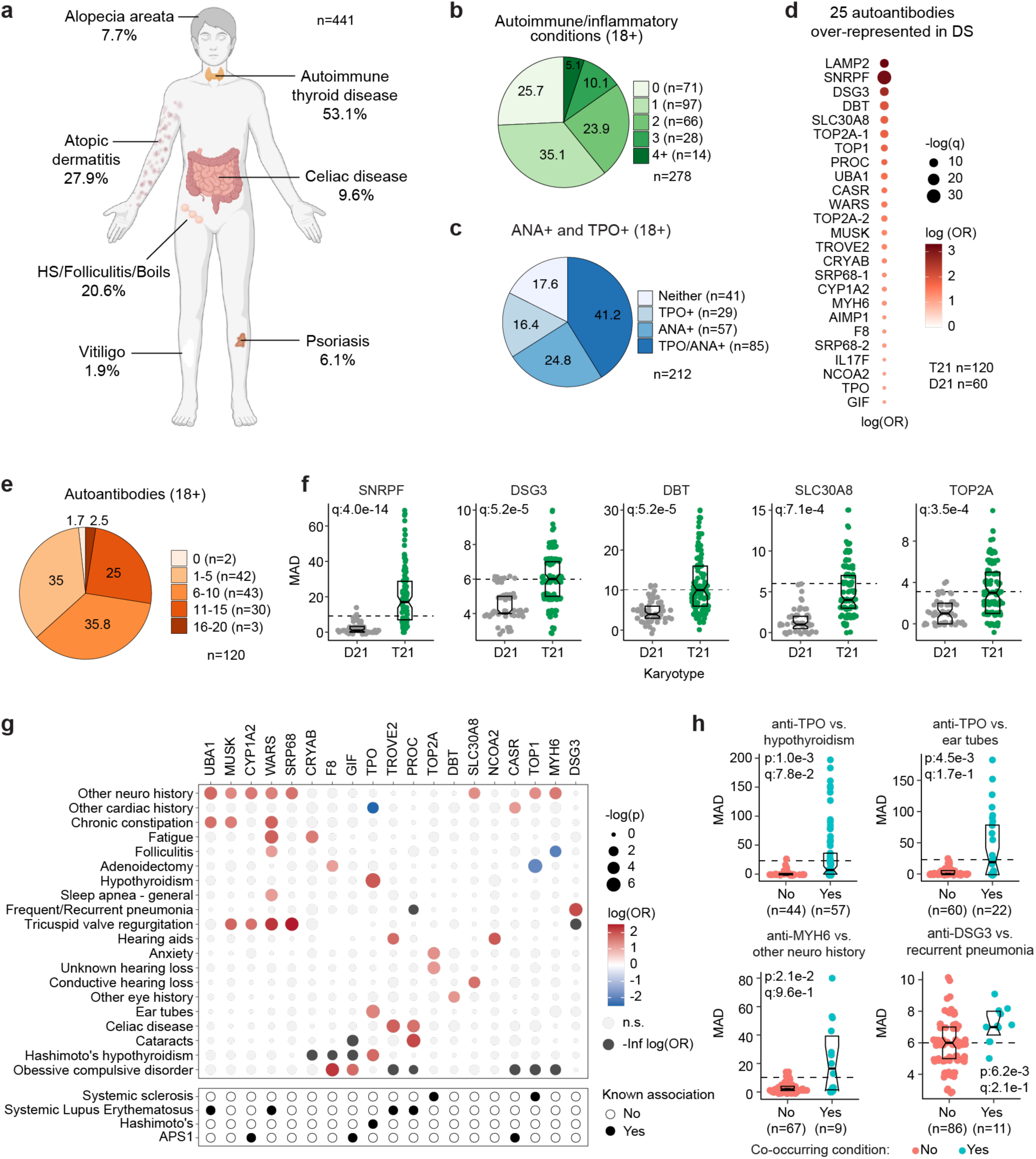
Multi-organ autoimmunity and widespread autoantibody production in Down syndrome. **a,** Overview of autoimmune and inflammatory conditions prevalent in persons with Down syndrome (DS) enrolled in the Human Trisome Project (HTP) cohort study. Percentages indicate the fraction of participants (n=441, all ages) with history of the indicated conditions. **b**, Pie chart showing autoimmune/inflammatory condition burden in adults (n=278, 18+ years old) with DS. **c**, Pie chart showing rates of positivity for anti-TPO and/or anti-nuclear antibodies (ANA) in adults (n=212, 18+ years old) with DS. **d**, Bubble plot displaying odds-ratios and significance for 25 autoantibodies with elevated rates of positivity in individuals with DS (n=120) versus 60 euploid controls (D21). q values calculated by Benjamini-Hochberg adjustment of p-values from Fisher’s exact test. **e**, Pie chart showing fractions of adults with DS (n=120, 18+ years old) testing positive for various numbers of the autoantibodies identified in d. **f**, Representative examples of autoantibodies more frequent in individuals with T21 (n=120) versus euploid controls (D21, n=60). MAD: median absolute deviation. Dashed lines indicate the positivity threshold of 90^th^ percentile for D21. Data are presented as modified sina plots with boxes indicating quartiles. **g**, Bubble plots showing the relationship between autoantibody positivity and history of various clinical diagnoses in DS (n=120). Size of bubbles is proportional to -log-transformed p values from Fisher’s exact test. **h**, Sina plots displaying the levels of selected autoantibodies in individuals with DS with or without the indicated co-occurring conditions. MAD: median absolute deviation. Dashed lines indicate the positivity threshold of 90^th^ percentile for D21. Sample sizes are indicated under each plot. q values calculated by Benjamini-Hochberg adjustment of p-values from Fisher’s exact tests.

**Figure 1 – figure supplement 1.**
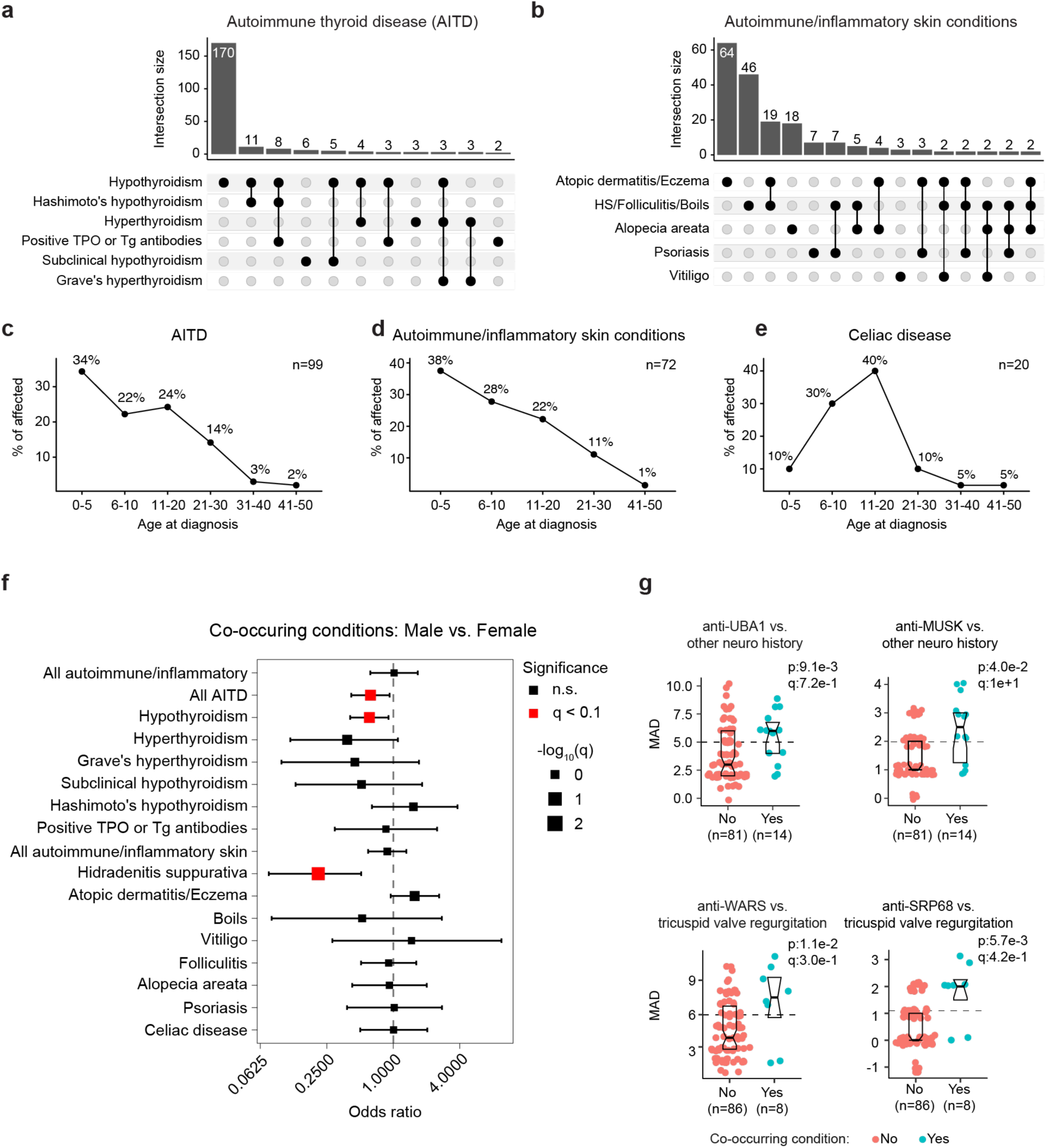
**Early onset multi-organ autoimmunity and autoantibody production in Down syndrome. a-b**, Upset plots showing overlap between various reported diagnoses indicative of autoimmune thyroid disease (a) or autoimmune/inflammatory skin conditions (b) in research participants with Down syndrome (DS, all ages, n=441) enrolled in the Human Trisome Project (HTP). **c-e,** Plots showing the percentages of cases by age at diagnosis for AITD (c), autoimmune/inflammatory skin conditions (d), and celiac disease (e). Sample sizes indicated in each chart. **f**, Odds ratio plot for Fisher’s exact test of proportions (cases vs. controls in males vs. females) for history of co-occurring conditions in individuals with DS (all ages, total n=441). Conditions with q < 0.1 (10% FDR) are highlighted in red. The size of square points is inversely proportional to q value; error bars represent 95% confidence intervals. **g**, Sina plots displaying the levels of select autoantibodies in individuals with DS, with or without history of the indicated co-occurring conditions. MAD: median absolute deviation. Horizontal dashed lines indicate 90th percentiles for the D21 group. Sample sizes are indicated under each plot. q values calculated by Benjamini-Hochberg adjustment of p-values from Fisher’s exact tests.

In the general population, the risk of autoimmune conditions increases with age and is higher in females, with autoimmune conditions tending to cluster, whereby occurrence of one autoimmune condition predisposes to a second condition^36,37^. Within the HTP cohort, analysis of age trajectories of immune-related conditions in DS revealed early onset, with >80% of AITD, autoimmune/inflammatory skin conditions, and celiac disease being diagnosed in the first two decades of life (**Figure 1 - supplement 1c-e**). The cumulative burden of autoimmunity and autoinflammation is similar in males versus females with DS, albeit with slightly increased rates of AITD and hidradenitis suppurativa in females (**Figure 1 - supplement 1f**). In terms of co-occurrence, when evaluating the adult population (18+ years old) for AITD, autoimmune/inflammatory skin conditions and celiac disease, we found that 75% of participants had a history of at least one condition, 38.4% had at least two, and 13.6% had three or more conditions (**Figure 1b**).

Interestingly, analysis of medical records found an unexpectedly low number of individuals with records of autoantibodies against the thyroid gland [4.3%, e.g., anti-thyroid peroxidase (TPO), anti-thyroglobulin (TG)] within the HTP cohort (**Figure 1 - supplement 1a**). This could be explained by the fact that thyroid disease is commonly diagnosed through measurements of thyroid-relevant hormones (TSH, T3, T4) without concurrent testing of autoantibodies. To investigate further, we measured anti-TPO levels as well as levels of anti-nuclear antibodies (ANA), a more general biomarker of autoimmunity (**Supplementary file 2**). Remarkably, 82.4% of adults with DS show positivity for at least one of these autoantibodies, with 41.2% being positive for both (**Figure 1c**). Indeed, 62% of individuals with history of hypothyroidism were TPO+, whereby anti-TPO is just one of the possible autoantibodies associated with AITD. Prompted by these results, we next completed a more comprehensive analysis of autoantibodies in DS using protein array technology, with a focus on ∼380 common autoepitopes from 270 proteins (see **Materials and Methods, Supplementary file 2**). These efforts identified 25 autoantibodies significantly over-represented in people with DS relative to age- and sex-matched controls (**Figure 1d**), with 98.3% of individuals with DS being positive for at least one of these autoantibodies, and 63.3% being positive for six or more (**Figure 1d-e**). In addition to autoantibodies against TPO, which is expressed exclusively in the thyroid gland, we identified autoantibodies targeting proteins that are either broadly expressed across multiple tissues (e.g., TOP1, UBA1, LAMP2) or preferentially expressed in specific organs across the human body, including liver (e.g., CYP1A2), pancreas (e.g., SLC30A8), skin (e.g., DSG3), bone marrow (e.g. SRP68), and brain tissue (e.g., AIMP1) (**Figure 1d, f**).

Analysis of autoantibody positivity relative to history of co-occurring conditions produced several interesting observations. Expectedly, individuals with hypothyroidism are more likely to be positive for anti-TPO antibodies (**Figure 1g-h**). However, unexpectedly, TPO+ status also associates with higher rates of use of pressure equalizing (PE) tubes employed to alleviate the symptoms of recurrent ear infections and otitis media with effusion (OME), which is common in DS^38^ (**Figure 1g-h**). Possible interpretations for this result are provided in the Discussion. Positivity for additional autoantibodies was more common in those with other co-occurring neurological conditions, a broad classification encompassing various seizure disorders, movement disorders, and structural brain abnormalities (**Figure 1g-h**, **Figure 1 – figure supplement 1g**). Salient examples are antibodies against MUSK, a muscle-associated receptor tyrosine kinase involved in clustering of the acetylcholine receptors in the neuromuscular junction^39^; UBA1, a ubiquitin conjugating enzyme involved in antigen presentation^40^; and MYH6, a cardiac myosin heavy chain isoform (**Figure 1g-h**, **Figure 1 – figure supplement 1g**). Individuals with history of tricuspid valve regurgitation display higher rates of four different autoantibodies, most prominently against WARS1, a tryptophan tRNA synthetase mutated in various neurodevelopmental disorders^41^, and SRP68, a protein commonly targeted by autoantibodies in necrotizing myopathies^42^ (**Figure 1 – figure supplement 1g**). Individuals with a history of frequent pneumonia present a higher frequency of autoantibodies against DSG3 (desmoglein 3), a cell adhesion molecule targeted by autoantibodies in paraneoplastic pemphigus (PNP), an autoimmune disease of the skin and mucous membranes that can involve fatal lung complications^43^ (**Figure 1g-h**).

Altogether, these results demonstrate widespread multi-organ autoimmunity across the lifespan in people with DS, with production of multiple autoantibodies that could potentially contribute to a number of co-occurring conditions more common in this population.

### Trisomy 21 causes global immune remodeling regardless of evident clinical autoimmunity

Several immune cell changes have been proposed to underlie the autoimmunity-prone state of DS^15,16,25,44^, but specific immune cell-to-phenotype associations have not been established in previous studies using smaller sample sizes. Therefore, we next investigated immune cell changes associated with various clinical and molecular markers of autoimmunity in DS. Toward this end we analyzed mass cytometry data from 292 individuals with DS relative to 96 euploid controls and tested for potential differences in immune cell subpopulations, identified using FlowSOM^45^, within the DS cohort based on number of autoimmune/inflammatory disease diagnoses, ANA positivity, TPO positivity, and positivity for additional autoantibodies. In agreement with previous analyses^15,16,25,44^, we observed massive immune remodeling in all major myeloid and lymphoid subsets, including increases in basophils, along with depletion of eosinophils and total B cells (**Figure 2a-c**, **Figure 2 – figure supplement 1a-c**). When comparing various subgroups within the DS cohort based on autoimmunity status, we observed that these global immune changes are largely independent of the presence of clinical diagnoses or autoantibody positivity, with very few additional changes significantly associated with these measures of autoimmunity (**Figure 2 – figure supplement 1d**). For example, the significant depletion of B cells and enrichment of basophils in DS is not significantly different among the various subgroups (**Figure 2c**, **Figure 2 – figure supplement 1c**). Among CD45+ CD66^lo^ non-granulocytes, most changes are conserved among subgroups, with the sole of exception of non-classical monocytes, which are further elevated in the ANA+ group (**Figure 2 – figure supplement 1d-e**). Among T cells, the overall pattern of depletion of naïve subsets and enrichment of differentiated subsets characteristic of DS^15,16,28,44^ is conserved across subgroups, as illustrated by consistent depletion of CD8+ naïve subsets along with increases in the CD8+ terminally differentiated effector memory (TEMRA) subset (**Figure 2f**, **Figure 2 – figure supplement 1d**). Notably, we observed depletion of ψ8 T cells (both total and CD8+) in those with multiple autoimmune diagnoses (**Figure 2 – figure supplement 1d, f**), a result that is in line with reports documenting depletion of these subsets from peripheral circulation toward sites of active autoimmunity^46^.

**Figure 2.**
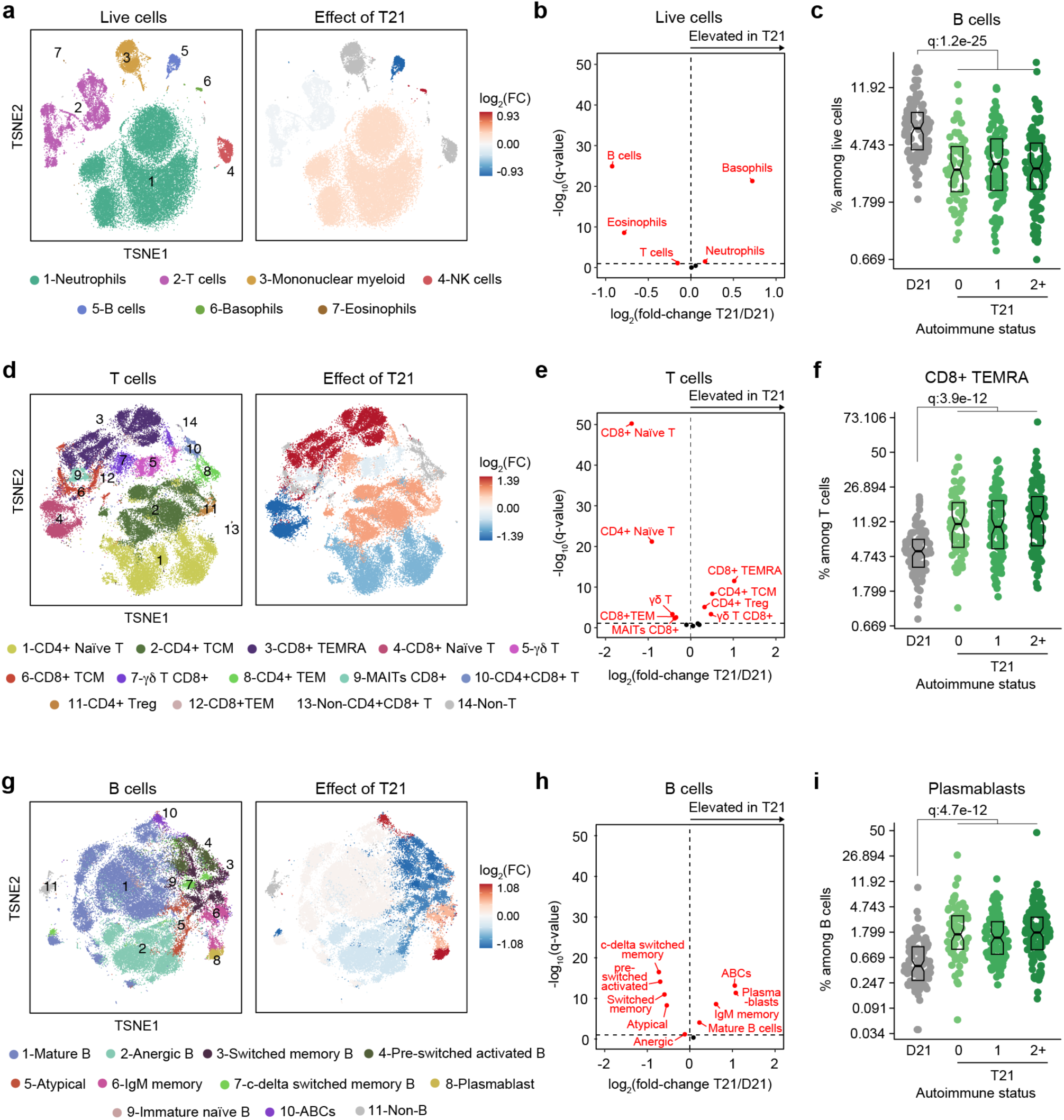
Trisomy 21 causes global immune remodeling regardless of clinically evident autoimmunity. **a,** t-distributed Stochastic Neighbor Embedding (t-SNE) plot displaying major immune populations identified by FlowSOM analysis of mass cytometry data for all live cells (left) and color coded by significant impact of T21 (beta regression q<0.1) on their relative frequency (right). Red indicates increased frequency and blue indicates decreased frequency among research participants with T21 (n=292) versus euploid controls (D21, n=96). **b**, Volcano plot showing the results of beta regression analysis of major immune cell populations among all live cells in research participants with T21 (n=292) versus euploid controls (D21, n=96). The dashed horizontal line indicates a significance threshold of 10% FDR (q<0.1) after Benjamini-Hochberg correction for multiple testing. **c**, Frequencies of B cells among all live cells in euploid controls (D21, n=96) versus individuals with T21 and history of 0 (n=69), 1 (n=102) or 2+ (n=121) autoimmune/inflammatory conditions. Data is displayed as modified sina plots with boxes indicating quartiles. **d-f**, Description as in a-c, but for subsets of T cells. **g-i**, Description as in a-c, but for subsets of B cells.

**Figure 2 – figure supplement 1.**
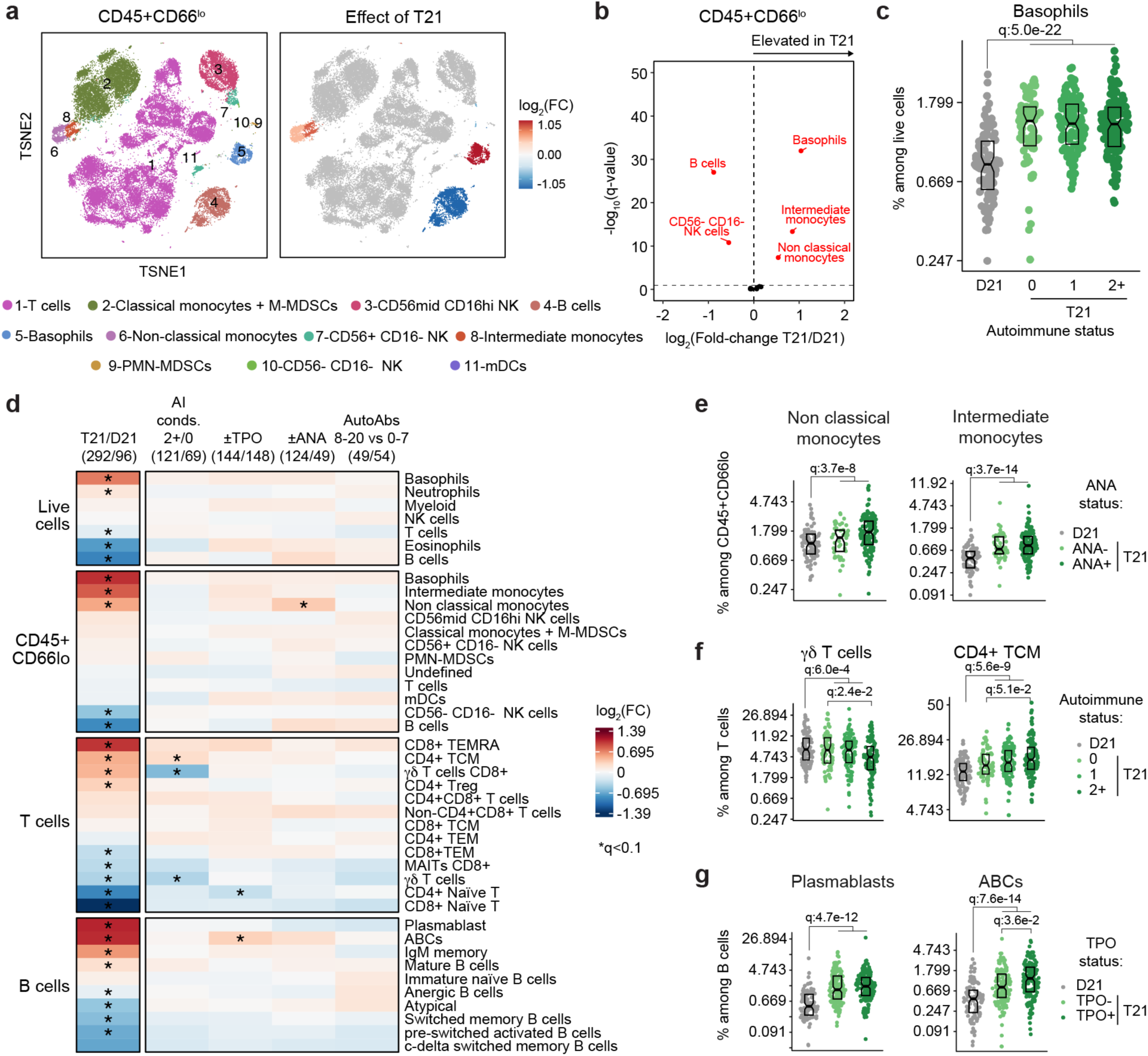
Consistent remodeling of the peripheral immune system in Down syndrome. **a,** t-distributed Stochastic Neighbor Embedding (t-SNE) plot displaying major immune populations identified by FlowSOM analysis of mass cytometry data for CD45+ CD66^lo^ non- granulocytes (left) and color coded by the impact of trisomy 21 (T21) on their relative frequency (right). Red indicates increased frequency and blue indicates decreased frequency among research participants with T21 (n=292) versus euploid controls (D21, n=96). **b**, Volcano plot showing the results of beta regression analysis of immune cell populations among CD45+ CD66^lo^ non-granulocytes from research participants with T21 (n=292) versus euploid controls (D21, n=96). The dashed horizontal line indicates a significance threshold of 10% FDR (q<0.1) after Benjamini-Hochberg correction for multiple testing. **c**, Frequencies of basophils among all live cells in euploid controls (D21, n=96) versus individuals with T21 and history of 0 (n=44), 1 (n=71) or 2+ (n=88) autoimmune/inflammatory conditions. Data is displayed as modified sina plots with boxes indicating quartiles. **d**, Heatmap summarizing the results of beta regression testing for differences in frequencies of indicated immune cell populations among all live cells, CD45+ CD66^lo^ non-granulocytes, T cells, and B cells by T21 (n=292) versus D21 (n=96) status, or by different subgroups within the T21 cohort: 2+ (n=88) versus 0 (n=44) autoimmune/inflammatory conditions; TPO+ (n=144) versus TPO- (n=148); ANA+ (n=124) versus ANA- (n=49); or positivity for 8-20 (n=49) versus 0-7 (n=54) autoantibodies elevated in DS. Asterisks indicate significance after Benjamini-Hochberg correction for multiple testing (q<0.1, 10% FDR). **e-g**, Representative examples of immune cell populations from d, showing effects of ANA positivity (e), number of autoimmune conditions (f), and TPO status (g). Data are presented as modified sina plots with boxes indicating quartiles, with q-values indicating beta regression significance after Benjamini- Hochberg correction for multiple testing.

We also observed slight elevation of CD4+ T central memory cells (TCM) (**Figure 2 – figure supplement 1d, f**). Among B cells, the overall shift toward more differentiated states such as plasmablasts, age- associated B cells (ABCs), and IgM+ memory cells is also conserved among subgroups, with the sole exception of ABCs, which tend to be further elevated in the TPO+ group (**Figure 2g-i**, **Figure 2 – figure supplement 1d, g**).

Altogether, these results indicate that T21 causes global remodeling of the immune system toward an autoimmunity-prone and pro-inflammatory state, prior to clinically evident autoimmunity, and dwarfing any additional effects associated with confirmed diagnoses of autoimmune/inflammatory conditions or common biomarkers of autoimmunity.

### Trisomy 21 causes hypercytokinemia from an early age independent of autoimmunity status

It is well established that individuals with DS display elevated levels of many inflammatory markers, including several interleukins, cytokines, and chemokines known to drive autoimmune conditions, such as IL-6 and TNF-α^18,28,44,47^. However, the interplay between hypercytokinemia, individual elevated cytokines, and development of autoimmune conditions in DS remains to be elucidated. Therefore, we analyzed data available from the HTP cohort for 54 inflammatory markers in plasma samples from 346 individuals with DS versus 131 euploid controls and cross-referenced these data with the presence of autoimmune conditions and autoantibodies. These efforts confirmed the notion of profound hypercytokinemia in DS^18,28,44,47^, with significant elevation of multiple acute phase proteins (e.g. CRP, SAA, IL1RA), pro-inflammatory cytokines (TSLP, IL-17C, IL-22, IL-17D, IL-9, IL-6, TNF-α) and chemokines (IP-10, MIP-3a, MIP-1a, MCP-1, MCP-4, Eotaxin), as well as growth factors associated with inflammation and wound healing (FGF, PlGF, VEGF-A) (**Figure 3a**). However, when evaluating for differences within the DS cohort based on various metrics of autoimmunity, we did not observe important differences based on number of autoimmune/inflammatory conditions, ANA or TPO positivity status, or number of other autoantibodies (**Figure 3a**). For example, CRP, IL-6, and TNF-α are equally elevated across all these subgroups (**Figure 3b-d**, **Figure 3 - figure supplement 1a**).

**Figure 3.**
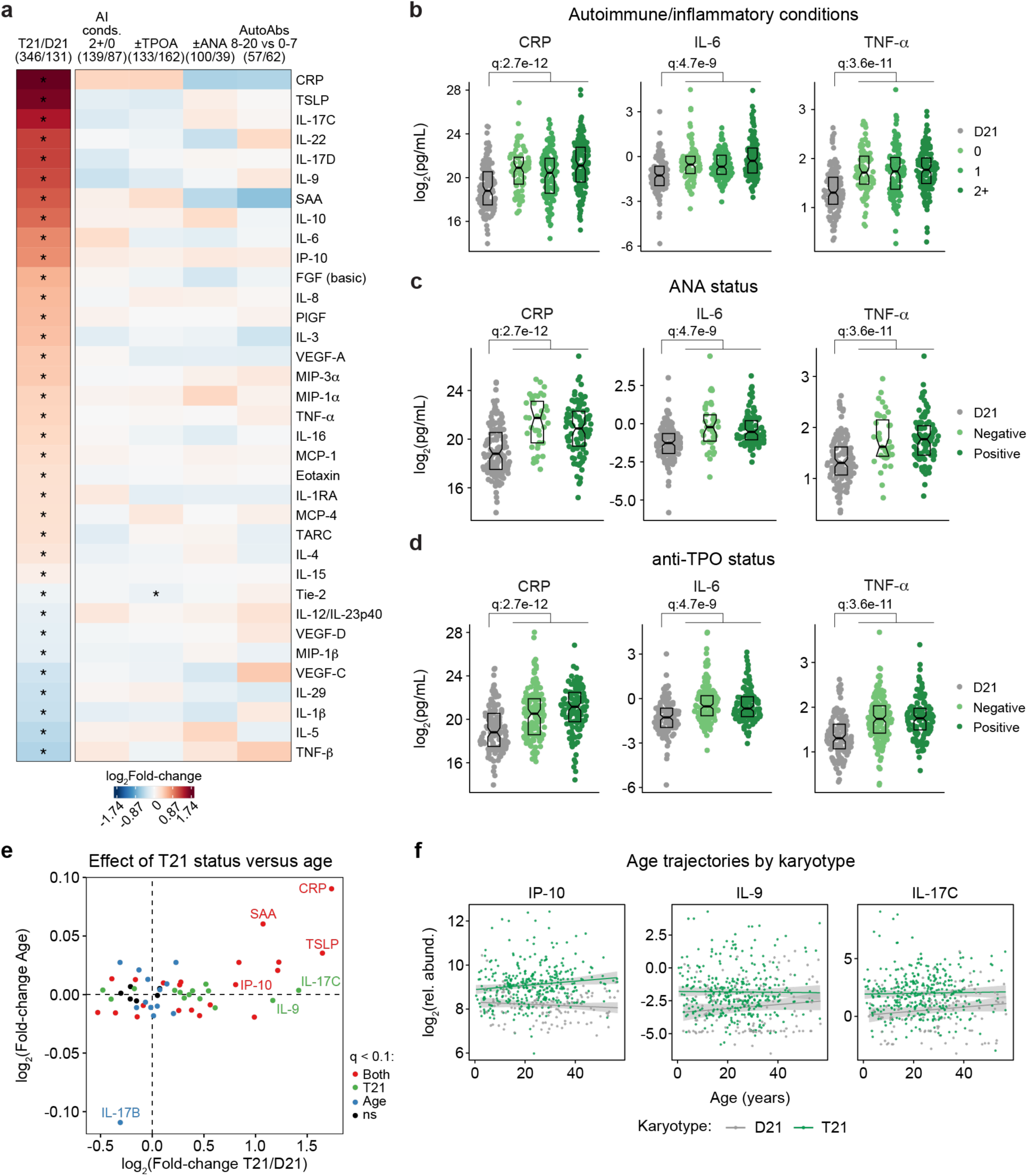
Trisomy 21 causes constitutive hypercytokinemia independent of autoimmunity status from an early age. **a,** Heatmap displaying log2-transformed fold-changes for plasma immune markers with significant differences in trisomy 21 (T21, n=346) versus euploid (D21, n=131), and between different subgroups within the T21 cohort: history of 2+ (n=139) versus 0 (n=87) autoimmune/inflammatory conditions (AI conds.); TPO+ (n=133) versus TPO- (n=162); ANA+ (n=100) versus ANA- (n=39); or positivity for 8-20 (n=57) versus 0-7 (n=62) autoantibodies (AutoAbs) elevated in DS. Asterisks indicate linear regression significance after Benjamini-Hochberg correction for multiple testing (q<0.1, 10% FDR). **b-d**, Comparison of CRP, IL-6 and TNF-α levels in euploid controls (D21, n=131) versus subsets of individuals with T21 based on number of autoimmune/inflammatory conditions (**b**), ANA positivity (**c**) or TPO positivity (**d**). Data are presented as modified sina plots with boxes indicating quartiles. Samples sizes as in a. q-values indicate linear regression significance after Benjamini-Hochberg correction for multiple testing. **e**, Scatter plot comparing the effect of T21 karyotype versus the effect of age in individuals with T21 (n=54 immune markers in 346 individuals with T21), highlighting immune markers that are significantly different by T21 status, age, or both. ns: not significantly different by T21 status or age. **f**, Scatter plots for example immune markers that are significantly elevated in T21, but which are either not elevated with age in the euploid (D21) cohort (i.e., IP-10), or in either the T21 (n=346) or D21 (n=131) cohorts. Lines represent least-squares linear fits with 95% confidence intervals in grey.

**Figure 3 – figure supplement 1.**
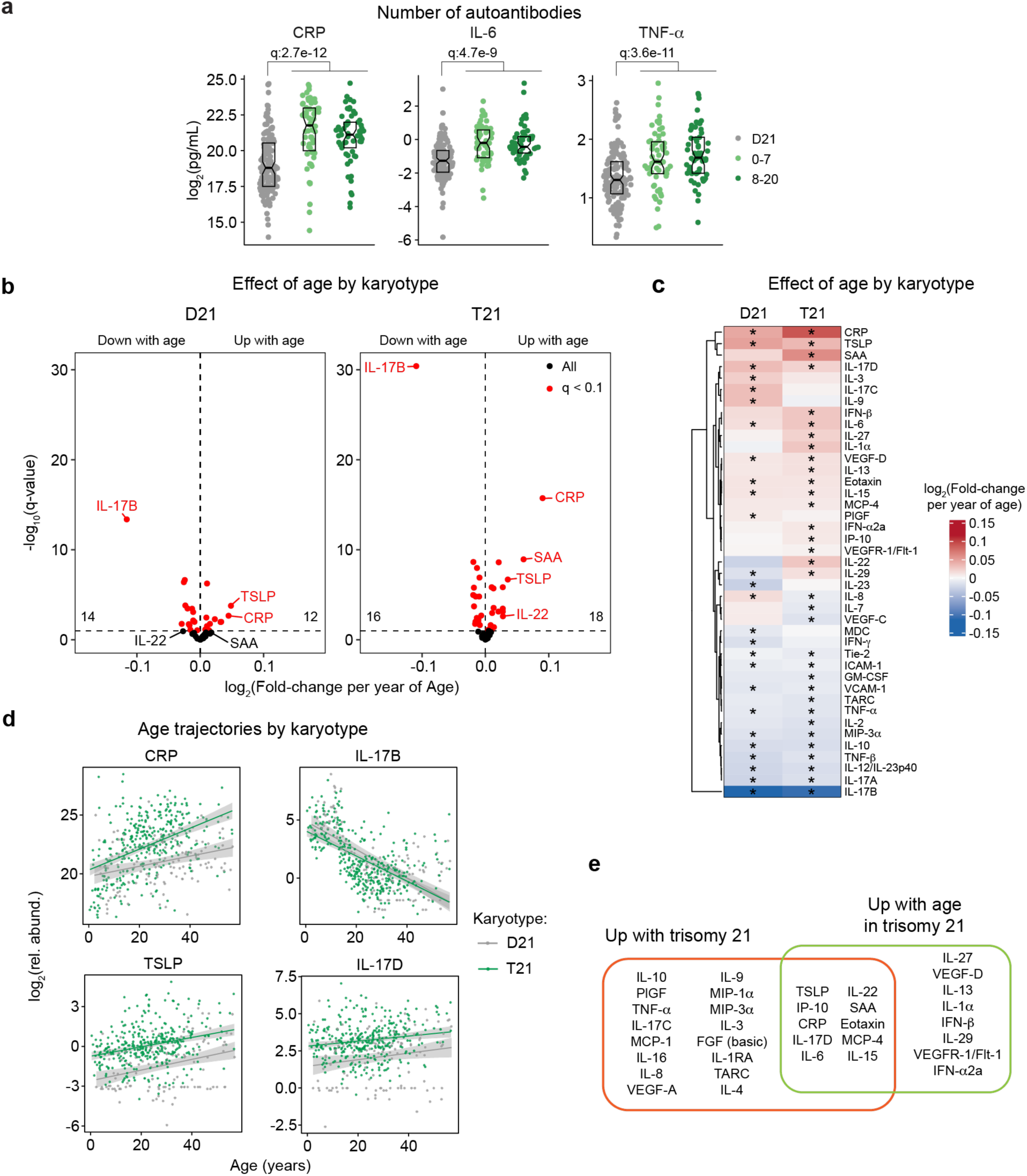
Consistent hypercytokinemia from an early age in Down syndrome. **a,** Comparison of CRP, IL-6, and TNF-α levels in euploid controls (D21, n=131) versus subsets of individuals with T21 based on number of autoantibodies commonly elevated in Down syndrome: 0-7 autoantibodies (n=62) versus 8-20 autoantibodies (n=57). Data are presented as modified sina plots with boxes indicating quartiles. q-values indicate linear regression significance after Benjamini-Hochberg correction for multiple testing. **b**, Volcano plots presenting the results of linear regression testing for association between age and the levels of 54 immune markers in the plasma of euploid controls (left, D21, n=131) and individuals with trisomy 21 (right, T21, n=346) enrolled in the Human Trisome Project (HTP) study. Horizontal dashed lines indicate a significance threshold of 10% FDR (q<0.1) after Benjamini-Hochberg correction for multiple testing. **c**, Heatmap comparing the effect of age on levels of immune markers in D21 and T21. Heatmap color scale represents log2-transformed mean fold-change per year of age; asterisks indicate significance (q < 0.1) for linear regression testing. **d**, Scatter plots showing the age trajectories of select immune markers in D21 versus T21. Sample sizes as in c. Lines represent least squares linear fits with shaded areas indicating 95% confidence interval. **e**, Diagram representing the overlap between immune markers elevated in T21 versus D21 and those elevated with age in T21.

Previous studies have reported signs of early immunosenescence and inflammaging in DS, including accelerated progression of immune lineages toward terminally differentiated states, early thymic atrophy, and elevated levels of pro-inflammatory markers associated with age in the typical population^25,27,48,49^. However, the extent to which the inflammatory profile of DS represents accelerated ageing versus other processes remains ill-defined. To address this, we first identified age-associated changes in immune markers within the euploid and DS cohorts separately (**Figure 3 - figure supplement 1b-c**). This exercise identified multiple immune markers that were up- or down-regulated with age, with an overall conserved pattern of age trajectories in both groups (**Figure 3 - figure supplement 1c**). For example, increased age is associated with increased CRP levels and decreased IL-17B levels in both cohorts (**Figure 3 – figure supplement 1d**). We then compared the effects of age versus T21 status on cytokine levels in the DS cohort, which identified many inflammatory factors elevated in DS across the lifespan that do not display a significant increase with age, such as IL-9 and IL-17C, or that increase with age only in the DS cohort, such as IP-10 (**Figure 3e-f**, **Figure 3 - figure supplement 1e**).

Altogether, these results indicate that T21 induces a constitutive hypercytokinemia from early childhood, with only a fraction of these inflammatory changes being exacerbated with age.

### A clinical trial for JAK inhibition in Down syndrome

Several lines of evidence support the notion that IFN hyperactivity and downstream JAK/STAT signaling are key drivers of immune dysregulation in DS^15–19,22,24,44,50^. In mouse models of DS, both normalization of *IFNR* gene copy number and pharmacologic JAK1 inhibition rescue their lethal immune hypersensitivity phenotypes^22,50^. Furthermore, we recently demonstrated that IFN transcriptional scores derived from peripheral immune cells correlate significantly with the degree of immune remodeling and hypercytokinemia in DS^44^, and we and others have reported the safe use of JAK inhibitors for treatment of diverse immune conditions in DS, including alopecia areata^51^, psoriatic arthritis^52^ and hemophagocytic lymphohistocytosis^53^ through small case series. Encouraged by these results, we launched a clinical trial to assess the safety and efficacy of the JAK inhibitor tofacitinib (Xeljanz, Pfizer) in DS, using moderate- to-severe autoimmune/inflammatory skin conditions as a qualifying criterion (NCT04246372). This trial is a single-site, open-label, Phase II clinical trial enrolling individuals with DS between the ages of 12 and 50 years old affected by alopecia areata, hidradenitis suppurativa, psoriasis, atopic dermatitis, or vitiligo (see qualifying disease scores in **Supplementary file 3**). After screening, qualifying participants are prescribed 5 mg of tofacitinib twice daily for 16 weeks, with an optional extension to 40 weeks (**Figure 4a**, see **Materials and Methods**). After enrollment and assessments at a baseline visit, participants attend five safety monitoring visits during the main 16-week trial period. The recruitment goal for this trial is 40 participants who complete 16 weeks of tofacitinib treatment, with a predefined IRB-approved qualitative interim analysis triggered when the first 10 participants completed the main 16-week trial (**Figure 4b**). Among the first 13 participants enrolled, one participant withdrew shortly after enrollment, one was excluded from analyses due to medication non-compliance (i.e., >15% missed doses), and one participant had not yet completed the trial at the time of the interim analysis (**Figure 4b**). Demographic characteristics of the 10 participants included in the interim analysis are shared in **Supplementary file 3**. Baseline qualifying conditions of the 10 participants included in the interim analysis were alopecia areata (n=6), hidradenitis suppurativa (n=3), and psoriasis (n=1) (open circles in **Figure 4c**). Two participants presented with concurrent atopic dermatitis, two with concurrent vitiligo, and two with concurrent hidradenitis suppurativa, albeit below the severity required to be the qualifying conditions (see closed circles in **Figure 4c**). In addition, seven participants had AITD/TPO+ and three had a celiac disease diagnosis (**Figure 4c**).

**Figure 4.**
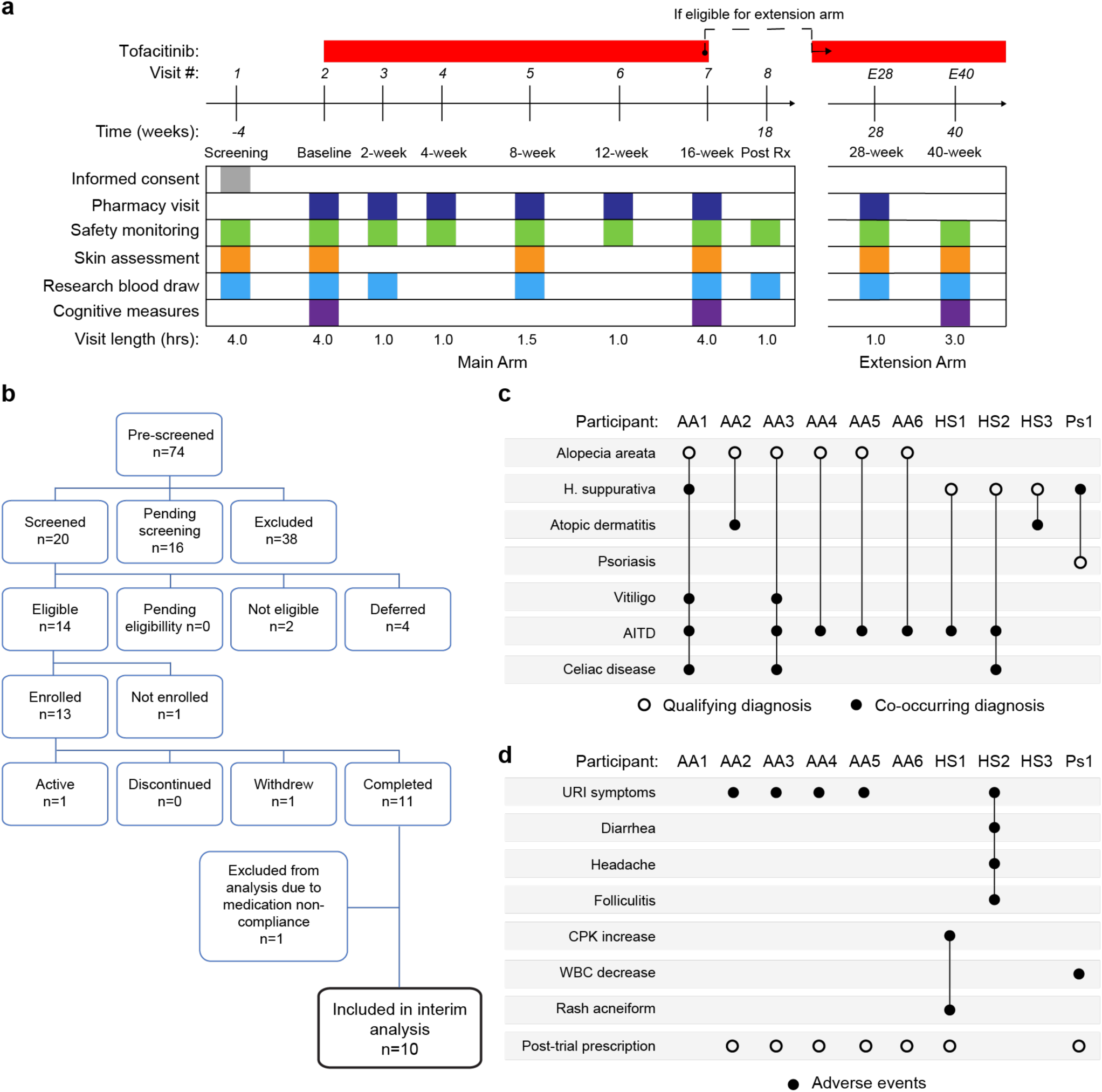
Clinical trial for JAK inhibition in Down syndrome. **a,** Schedule of activities for clinical trial of JAK inhibition in Down syndrome (NCT04246372). **b,** Consort chart for first 13 participants enrolled in the clinical trial. **c,** Upset plot displaying the qualifying and co-occurring autoimmune/inflammatory conditions for the 10 participants included in the interim analysis. **d,** Upset plots summarizing the adverse events annotated for the first 10 participants over a 16-week treatment period.

### Tofacitinib is well tolerated in Down syndrome

Analysis of adverse events (AEs) recorded for the 10 first participants over 16 weeks did not identify any AEs considered definitely related to tofacitinib treatment or classified as severe. Several AEs were annotated as ‘possibly related’ to treatment (**Figure 4d, Supplementary file 4**). Five episodes of upper respiratory infections (URIs) affecting five different participants were observed. Based on the safety data for tofacitinib in the general population^54^, all episodes of URIs were annotated as possibly related to treatment. Participant AA2 developed occasional cough and rhinorrhea that resolved with over-the- counter medication. Two other participants reported transient rhinorrhea (AA3, AA5). Participant AA4 developed a nasal congestion, with chest pain and a productive cough. This participant tested negative for SARS-Co-V2, Flu A-B, and RSV. Tofacitinib was not paused during this episode, and symptoms resolved with over-the-counter medication. Participant HS2 experienced a sore throat with middle ear inflammation that resolved with over-the-counter treatment. This participant also presented with folliculitis, which resolved with antibiotic treatment. Participant HS1 experienced a short transient elevation (<3 days) in creatine phosphokinase (CPK) that resolved spontaneously, and rash acneiform. Participant Ps1 experienced a transient and asymptomatic decrease in white blood cell (WBC) counts that resolved by the end of the trial.

Overall, tofacitinib treatment was not discontinued for any of the 10 participants over the 16-week study period, and seven participants eventually obtained off-label prescriptions after completing the trial and are currently taking the medicine. Based on these interim results, recruitment resumed and is ongoing.

### Tofacitinib improves diverse autoimmune/inflammatory skin conditions in Down syndrome

In the clinical trial, skin pathology is monitored using global metrics of skin health, including the Investigator’s Global Assessment (IGA) and the Dermatology Life Quality Index (DLQI), as well as disease-specific scores, such as the severity of alopecia tool (SALT), the psoriasis area and severity index (PASI), or the eczema area and severity index (EASI) (see **Materials and Methods, Supplementary file 5**. The interim analysis showed that seven of the ten participants had an improvement in the IGA score and eight of the ten reported some improvement on their life quality related to their skin condition as measured by the DLQI (**Figure 5a-b**). The most striking effects were observed for alopecia areata (**Figure 5c-d**, **Figure 5 – figure supplement 1a**). Five of six participants with alopecia areata showed scalp hair regrowth, with the exception being a male participant (AA1) with history of alopecia totalis for 20+ years who only showed facial hair and eyelash re-growth. One participant presented with psoriasis due to psoriatic arthritis and experienced an almost complete remission of psoriatic arthritis symptoms (Ps1, **Figure 5 – figure supplement 1b-c**). For the two participants that presented with atopic dermatitis, the clinical manifestations were markedly reduced during tofacitinib treatment (**Figure 5e-f**). A total of five participants were affected by HS, three of them as the qualifying condition (HS1-3). No clear trend was seen in the Modified Sartorius Scale (MSS) score used to monitor HS (**Figure 5 – figure supplement 1d-e**).

**Figure 5.**
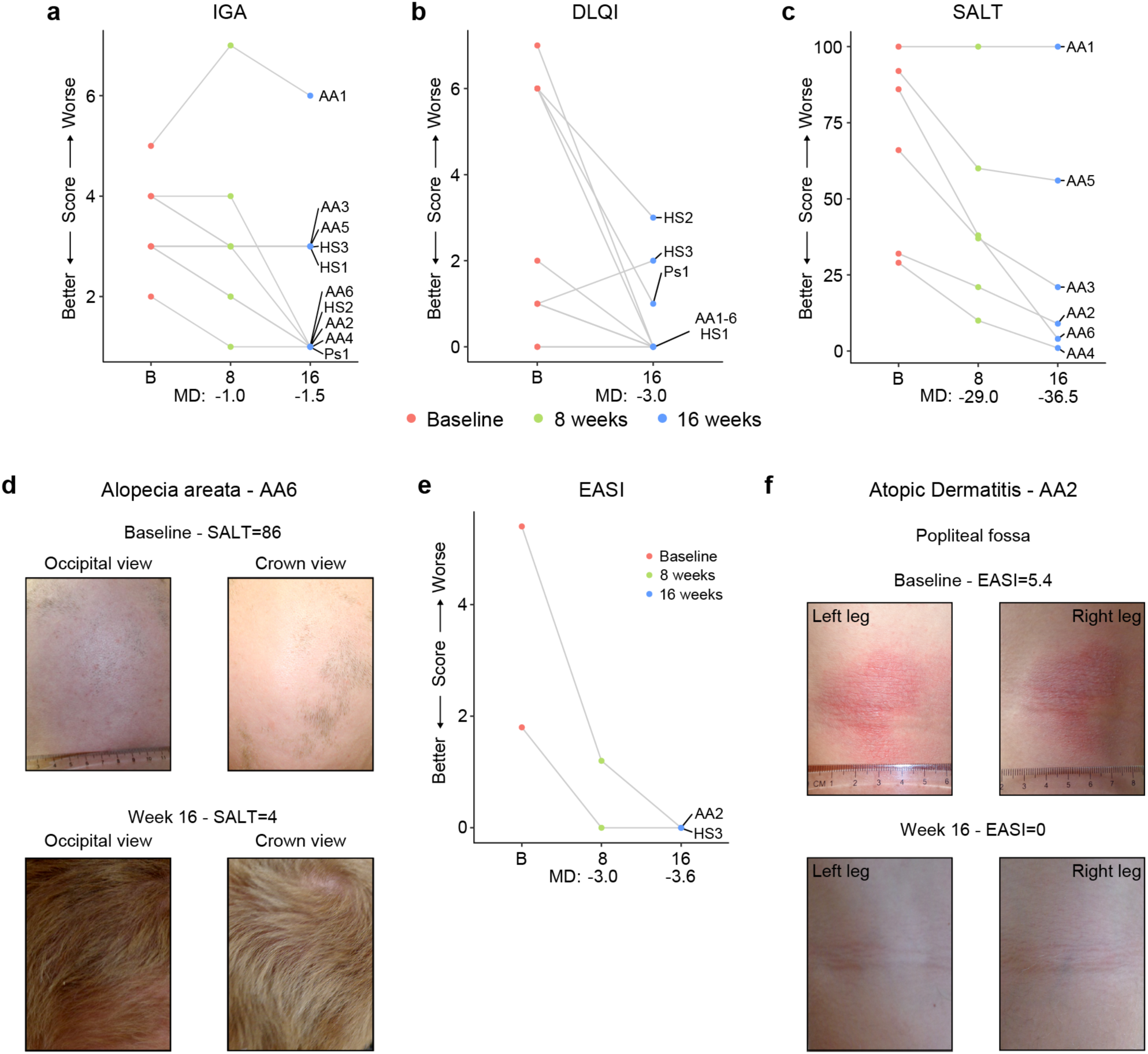
Tofacitinib improves diverse immune skin pathologies in Down syndrome. a-b, Investigator global assessment (IGA) scores (a) and Dermatological Life Quality Index (DLQI) scores (b) for the first 10 participants at baseline visit (B), mid-point (8 weeks) and endpoint (16 weeks) visits. MD: median difference. **c,** Severity of Alopecia Tool (SALT) scores for the first seven participants with alopecia areata in the trial. **d,** Images of participant AA6 at baseline versus week 16. **e,** Eczema Area and Severity Index (EASI) scores for two participants with mild atopic dermatitis. **f,** Images of participant AA2 showing improvement in atopic dermatitis upon tofacitinib treatment. p values not shown as per interim analysis plan.

**Figure 5 – figure supplement 1.**
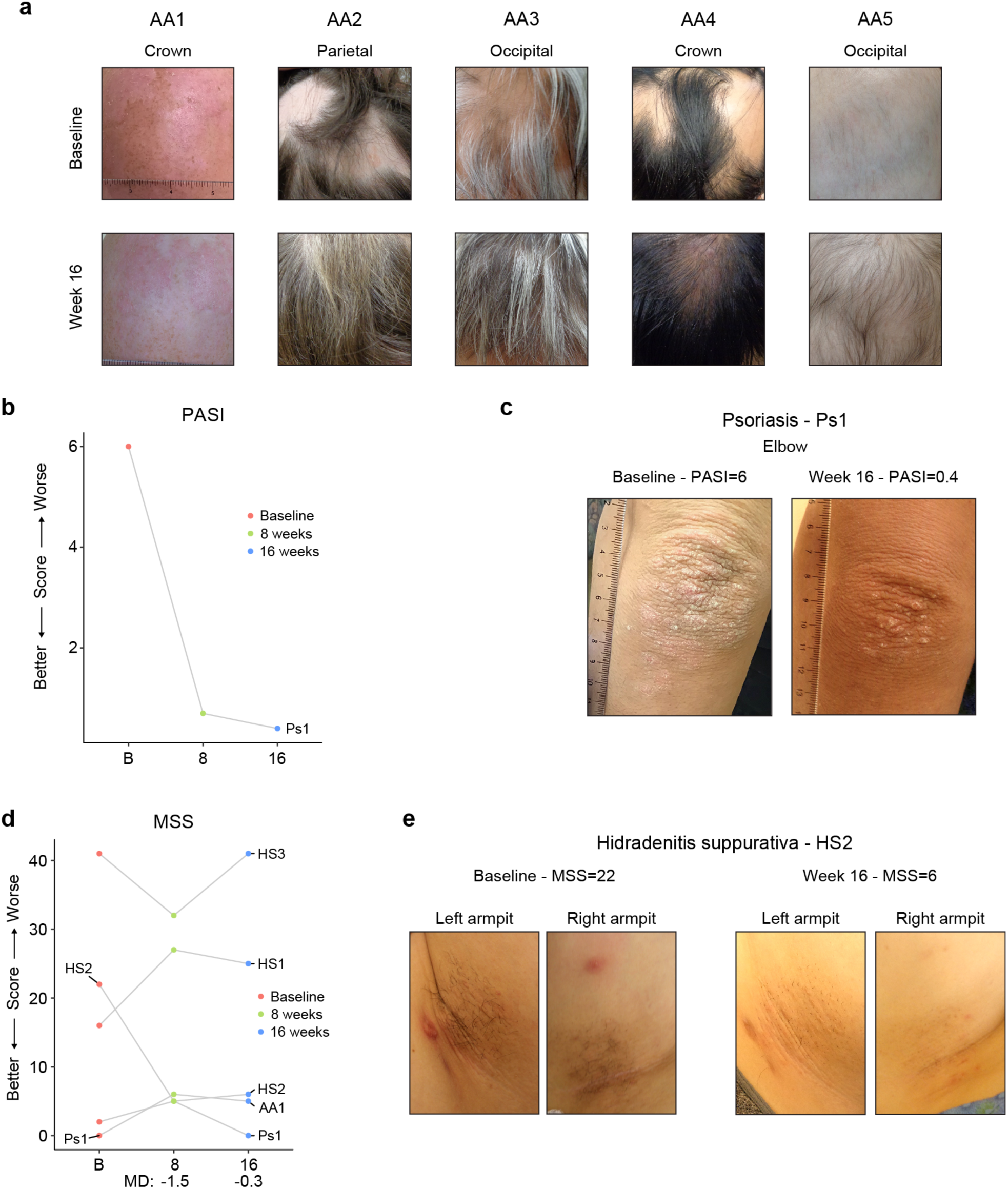
Tofacitinib improves diverse skin pathologies in Down syndrome. **a,** Images of five participants with alopecia areata at baseline and after 16 weeks of tofacitinib treatment. **b-c**, Psoriasis Area and Severity Index score (b) and images (c) for participant with psoriatic arthritis. **d,** Modified Sartorius Scale (MSS) scores for five participants with hidradenitis suppurativa (HS). MD: median difference. **e**, Images for participant affected by HS at baseline and 16-week endpoint visit. p values not shown as per interim analysis plan.

Altogether, these results indicate that JAK inhibition could provide therapeutic benefit for several autoimmune/inflammatory skin conditions more common in DS.

### Tofacitinib normalizes IFN scores and decreases pathogenic cytokines and autoantibodies

It is well demonstrated that individuals with DS display elevated IFN signaling across multiple immune and non-immune cell types^15–17,19^. Using an IFN transcriptional score composed of 16 interferon-stimulated genes (ISGs)^55^ measured via bulk RNA sequencing of peripheral blood RNA, individuals with DS in the HTP cohort study show a significant increase in these scores^50^ (**Figure 6a**). Reduction of IFN scores is designated as a primary endpoint in the trial. At baseline, clinical trial participants show IFN scores within the typical range for DS, but values are decreased at 2, 8, and 16 weeks of tofacitinib treatment (**Figure 6a, Supplementary file 6**). Time course analysis revealed that most participants show a decrease in IFN scores as soon as two weeks of treatment which is sustained over time, with two clear exceptions (**Figure 6 – figure supplement 1a**). At the 8-week study midpoint, 9 of 10 participants had decreased IFN scores relative to baseline, except participant AA2 who reported a COVID-19 vaccination three days prior to the visit and was pausing tofacitinib at the time of the blood draw (**Figure 6 – figure supplement 1a**). At the 16-week time point, nine of ten participants had decreased IFN scores, with the exception being AA4, who developed an URI in the week prior to the blood draw (**Figure 6 – figure supplement 1a**). Therefore, although all participants displayed decreased IFN scores at one or more time points during the treatment, IFN scores could be sensitive to immune triggers. Analysis of individual ISGs composing the IFN score revealed that whereas many ISGs elevated in DS display reduced expression upon tofacitinib treatment (e.g., *RSAD2*, *IFI44L*), others do not (e.g., *BPGM*) (**Figure 6b**, **Figure 6 – figure supplement 1b**). To investigate this further, we defined the impact of tofacitinib on all 136 ISGs significantly elevated in DS that are not encoded on chr21^44^ (**Figure 6c**). Collectively, ISGs as a group are significantly downregulated upon tofacitinib treatment, but the effect is not uniform across all ISGs (**Figure 6c**), indicating that JAK1/3 inhibition does not reduce all IFN signaling elevated in DS, which could be explained by the fact that the IFN pathways also employ JAK2 for signal transduction^56,57^. Global analysis of transcriptome changes revealed that tofacitinib treatment reverses the dysregulation of many gene signatures observed in DS, effectively attenuating many pro-inflammatory signatures beyond IFN gamma and alpha responses, such as Inflammatory Response, TNF-α signaling via NFkB, IL-2 STAT5 signaling, and IL-6 JAK STAT3 signaling (**Figure 6 – figure supplement 1c**). Tofacitinib also reversed elevation of genes involved in Oxidative Phosphorylation and dampened downregulation of gene sets involved in Wnt/Beta Catenin and Hedgehog Signaling (**Figure 6 – figure supplement 1c-d**). Conversely, tofacitinib did not rescue elevation of genes involved in Heme Metabolism or Mitotic Spindle (**Figure 6 – figure supplement 1c-d**), suggesting that these transcriptome changes are not tied to the inflammatory profile of DS.

**Figure 6.**
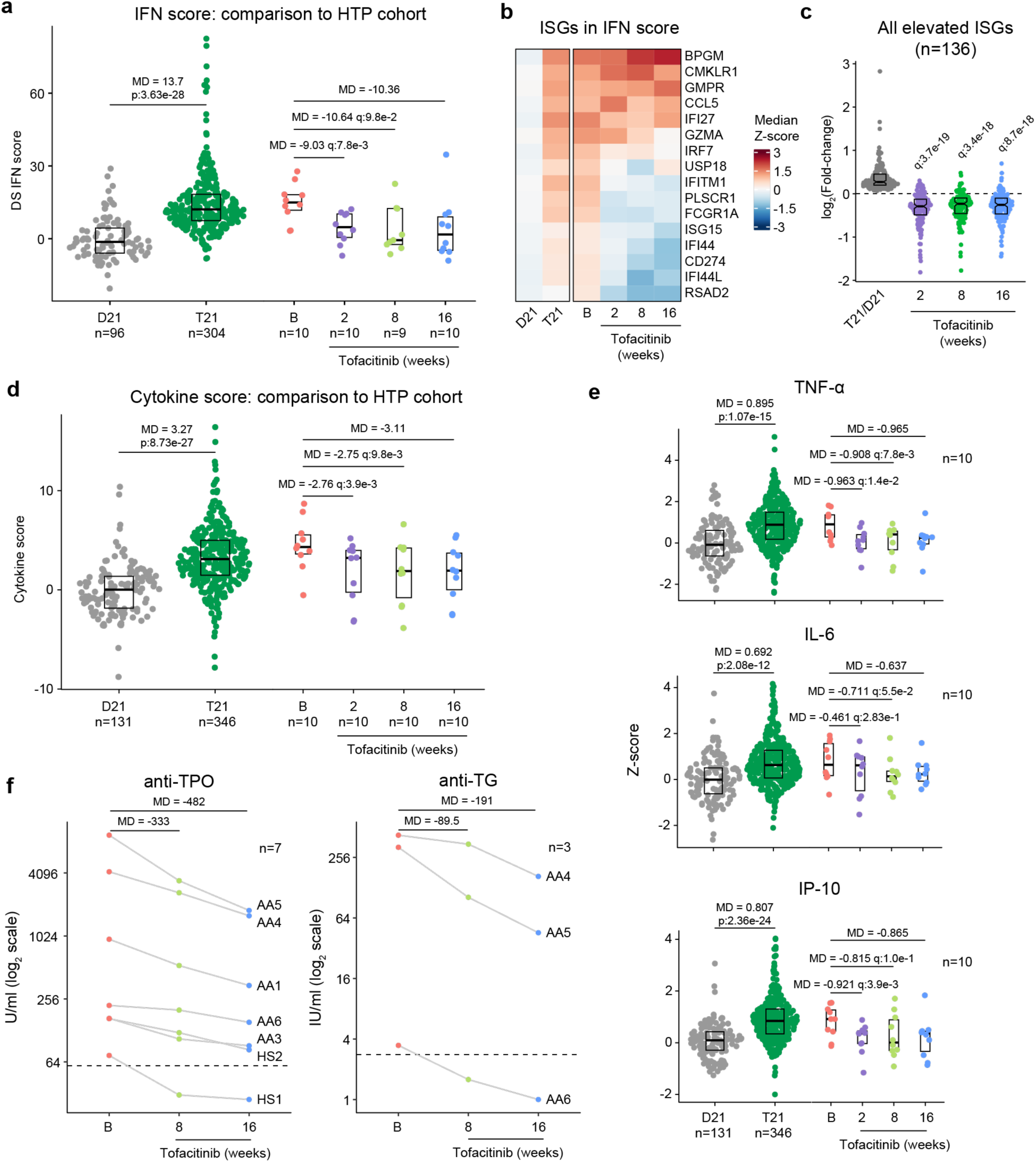
Tofacitinib reduces IFN scores, hypercytokinemia, and pathogenic autoantibodies in Down syndrome. **a,** Comparison of interferon (IFN) transcriptional scores derived from whole blood transcriptome data for research participants in the Human Trisome Project (HTP) cohort study by karyotype status (D21, grey; T21, green) and the clinical trial cohort at baseline (B), and weeks 2, 8 and 16 of tofacitinib treatment. Data are represented as modified sina plots with boxes indicating quartiles. Sample sizes are indicated below the x-axis. Horizontal bars indicate comparisons between groups with median differences (MD) with p-values from Mann-Whitney U-tests (HTP cohort) or q-values from paired Wilcox tests (clinical trial). q value for the 16-week endpoint is not shown as per interim analysis plan. **b,** Heatmap displaying median z-scores for the indicated groups (as in a) for the 16 interferon-stimulated genes (ISGs) used to calculate IFN scores. **c,** Analysis of fold changes for 136 ISGs not encoded on chr21 that are significantly elevated in Down syndrome (T21 versus D21) at 2, 8 and 16 weeks of tofacitinib treatment relative to baseline. Sample sizes as in a. q-values above each group indicate significance of Mann-Whitney U-tests against log2-transformed fold-change of 0 (no-chance), after Benjamini-Hochberg correction for multiple testing. **d,** Comparison of cytokine score distributions for the HTP cohort by karyotype status (D21, T21) versus the clinical trial cohort at baseline (B) and 2, 8 and 16 weeks of tofacitinib treatment. Data are represented as modified sina plots with boxes indicating quartiles. Sample sizes are indicated below the x-axis. Horizontal bars indicate comparisons between groups with median differences (MD) with p-values from Mann-Whitney U-tests (HTP cohort) and q-values from paired Wilcox tests (clinical trial). q value for the 16-week endpoint is not shown as per interim analysis plan. **e,** Comparison of plasma levels of cytokines in the HTP cohort by karyotype status (D21, T21) and the clinical trial cohort at baseline (B) versus 2, 8 and 16 weeks of tofacitinib treatment. Data are represented as modified sina plots with boxes indicating quartiles. Sample sizes are indicated below x-axis. Horizontal bars indicate comparisons between groups with median differences (MD) with p-values from Mann- Whitney U-tests (HTP cohort) and q values from paired Wilcox tests (clinical trial). q value for the 16- week endpoint is not shown as per interim analysis plan. **f,** Plots showing levels of autoantibodies against thyroid peroxidase (TPO) and thyroglobulin (TG) at baseline versus 8 and 16 weeks of tofacitinib treatment. Sample sizes are indicated in each plot.

**Figure 6 – figure supplement 1.**
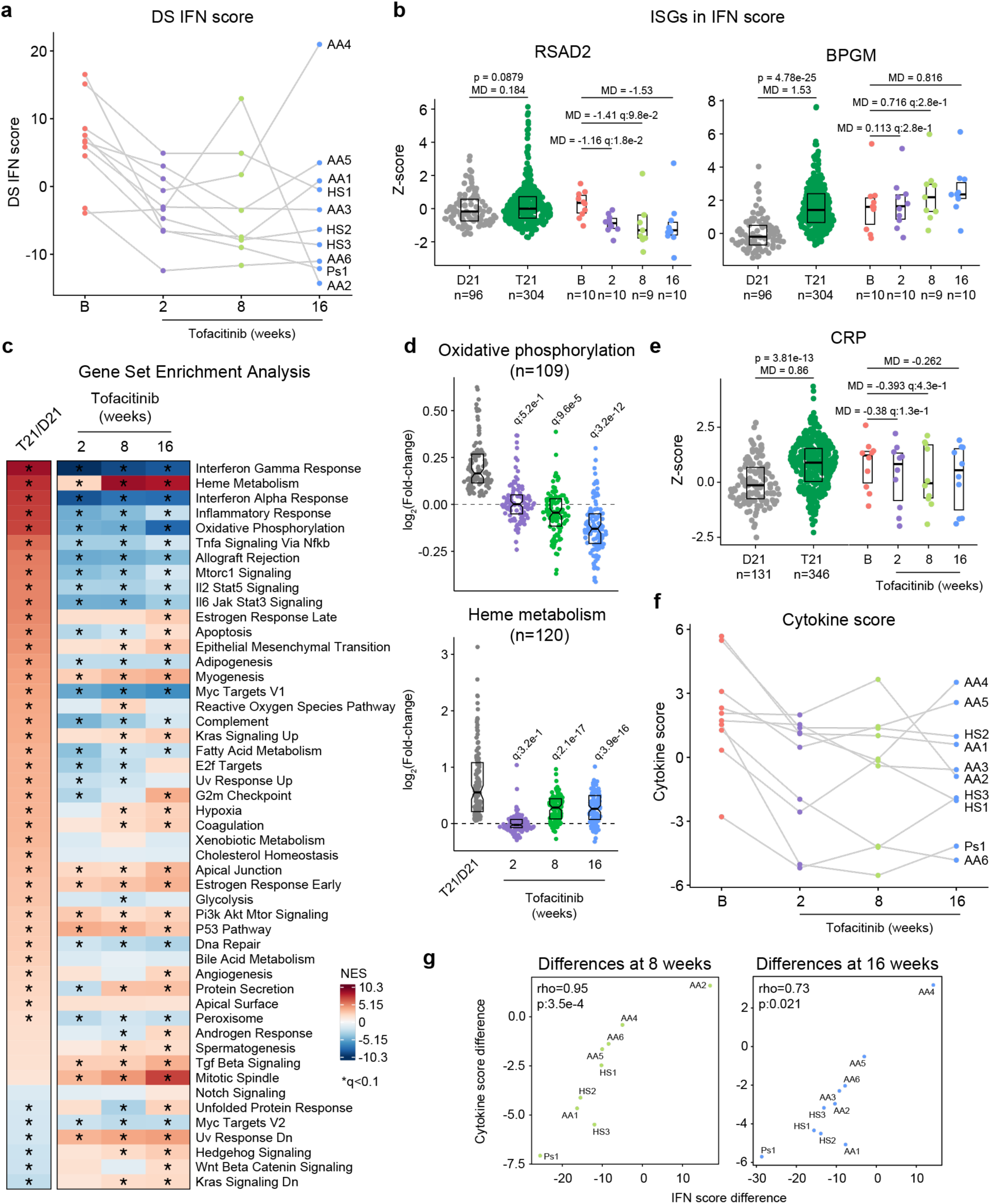
JAK inhibition reduces multiple markers of inflammation and autoimmunity in Down syndrome. **a,** Plot showing trajectory of IFN scores derived from whole blood transcriptome for 10 clinical trial participants at baseline (B), versus 2, 8 and 16 weeks of tofacitinib treatment. **b**, Comparison of ISG expression in the whole blood transcriptome data from research participants in the Human Trisome Project (HTP) cohort study by karyotype status (D21, grey; T21, green) and the clinical trial cohort at baseline (B), and weeks 2, 8 and 16 of tofacitinib treatment. Data are represented as modified sina plots with boxes indicating quartiles. Sample sizes are indicated below x- axis. Horizontal bars indicate comparisons between groups with median differences (MD) with p-values from Mann-Whitney U-tests (HTP cohort) and q-values from paired Wilcox tests (clinical trial). **c,** Heatmap displaying the results of Gene Set Enrichment Analysis (GSEA) of global transcriptome changes in the whole blood RNA of research participants in the HTP cohort (T21, n=304; D21, n=96) versus the clinical trial cohort at 2 (n=10), 8 (n=9), and 16 weeks (n=10) of tofacitinib treatment relative to baseline (n=10). Asterisks indicate significance after correction by Benjamini-Hochberg method for multiple testing (q<0.1, 10% FDR). NES: normalized enrichment score. **d,** Analysis of fold changes for 109 genes involved in oxidative phosphorylation and 120 genes involved in heme metabolism significantly elevated in Down syndrome (T21 versus D21 in the HTP cohort) versus the clinical trial cohort at 2, 8 and 16 weeks of tofacitinib treatment relative to baseline. Sample numbers as in c. **e,** Comparison of CRP levels in the HTP cohort by karyotype status (D21, grey; T21, green) versus the clinical trial cohort at baseline (B) and 2, 8 and 16 weeks of tofacitinib treatment. Data are represented as modified sina plots with boxes indicating quartiles. Sample sizes are indicated below x-axis. Horizontal bars indicate comparisons between groups with median differences (MD) with p-values from Mann-Whitney U-tests (HTP cohort) and q-values from paired Wilcox tests (clinical trial). **f,** Plot showing trajectory of cytokine scores for 10 clinical trial participants at baseline (B), versus 2, 8 and 16 weeks of tofacitinib treatment. **g,** Plots showing Spearman correlations between fold changes in IFN scores versus cytokine scores at 8 and 16 weeks of tofacitinib treatment versus baseline. Sample size is n=10.

A secondary endpoint in the trial is decrease of peripheral inflammatory markers as defined by a composite cytokine score derived from measurements of TNF-α, IL-6, CRP, and IP-10, and which is significantly increased in participants with DS in the HTP study (**Figure 6d**). At baseline, clinical trial participants show cytokine scores within the range observed for DS, but these values decrease at 2, 8 and 16 weeks relative to baseline (**Figure 6d-e**, **Figure 6 – figure supplement 1e**). The decreases in TNF-α and IL-6 observed upon tofacitinib treatment indicates that elevation of these potent inflammatory cytokines requires sustained JAK/STAT signaling in DS (**Figure 6e**). As for the IFN scores assessment, time course analysis revealed that most participants show decreases in cytokine scores within two weeks of treatment that are sustained over time, again with the exception of AA2 at week 8 and AA4 at week 16. This reveals a correspondence between RNA-based transcriptional IFN scores and circulating levels of these cytokines in plasma, while also illustrating that both metrics may remain sensitive to immune triggers (**Figure 6 – figure supplement 1f-g**).

One tertiary endpoint of the trial investigates the impact of tofacitinib treatment on levels of autoantibodies and markers employed to diagnose AITD [e.g., anti-TPO, anti-TG, anti-thyroid stimulating hormone receptor (TSHR)] and celiac disease [e.g., anti-tissue transglutaminase (tTG), anti-deamidated gliadin peptide (DGP)]. Seven of the 10 participants presented at baseline with anti-TPO levels above the upper limit of normal (ULN, 60U/mL), and all seven experienced a decrease in these auto-antibodies at 8 weeks and 16 weeks relative to baseline (**Figure 6f**). In fact, for one participant (HS1) the levels decreased below the ULN while on the trial. All seven of these participants had a history of thyroid disease (**Figure 4c**), which was being medically managed and/or clinically monitored with acceptable TSH and T4 values. Additionally, three of these seven participants also had anti-TG levels above the ULN (4 IU/mL) and all three showed a decrease from baseline levels while on tofacitinib at both 8 and 16 weeks, with one participant (AA6) falling below the ULN upon treatment (**Figure 6f**). Three participants also had anti- TSHr levels above the ULN, but no clear changes were observed upon treatment (**Supplementary file 5**). None of the 10 participants displayed anti-tTG or anti-DGP levels detected above ULN at screening.

Altogether, these results indicate that tofacitinib treatment decreases IFN scores, levels of key pathogenic cytokines, and key autoantibodies involved in AITD. Importantly, tofacitinib treatment lowers IFN scores and cytokine levels to within the range observed in the general population, not below, indicating that this immunomodulatory strategy can provide therapeutic benefit in DS without overt immune suppression.

## Discussion

An increasing body of evidence indicates that immune dysregulation contributes to the pathophysiology of DS and that immunomodulatory therapies could provide multidimensional benefits in this population. In mouse models, triplication of four *IFNR* genes contributes to multiple hallmarks of DS^50,58^ and JAK inhibition attenuates global dysregulation of gene expression^44^ while rescuing key phenotypes, such as lethal immune hypersensitivity^22^ and CHDs^24^. The fact that gene signatures of IFN hyperactivity are present in human embryonic tissues with T21^59^ and embryonic tissues from mouse models of DS^50,60^ indicates that the harmful effects of IFN hyperactivity could start *in utero*, supporting the notion that DS could be understood, in part, as an inborn error of immunity with similarities to monogenic interferonopathies^61^.

Results presented here demonstrate that T21 causes widespread multi-organ autoimmunity of pediatric onset, with production of autoantibodies targeting every major organ system. These results justify additional efforts to define the key pathogenic autoantibodies in DS beyond those commonly associated with AITD and celiac disease. Our analysis found significant associations between specific autoantibodies and some conditions more common in DS, but the diagnostic value of these observations will require validation efforts in much larger cohorts, which could lead to a personalized medicine approach for the management of autoimmunity in DS. For example, we found autoantibodies associated with various forms of auditory dysfunction (**Figure 1g**), suggesting the possibility of autoimmune hearing loss in DS^62^. Elevated levels of anti-TPO in individuals with history of use of ear tubes suggests an interplay between otitis media and endocrine dysfunction in DS^63^. For example, it is possible that recurrent ear infections cause a chronic immune stimulus that lead to eventual breach of tolerance in this autoimmunity-prone population, even perhaps through epitope mimicry^64^. Antibodies targeting MUSK, which we found to be elevated in DS and associated with co-occurring neurological phenotypes (**Figure 1g-h**), have been linked to development of myasthenia gravis, a chronic autoimmune neuromuscular disease that causes weakness in the skeletal muscles^65^. Whether MUSK antibodies associate with similar phenotypes in DS will require further investigation. Elevation of SRP68 autoantibodies in DS (**Figure 1d,f**), which are common in necrotizing myopathies with cardiovascular involvement^42^, suggests a potential autoimmune basis for musculoskeletal and cardiovascular complications in DS, which also warrants additional research.

We observed constitutive global immune remodeling and hypercytokinemia regardless of reported diagnoses of autoimmune disease or measurable autoantibody production from an early age, indicative of an autoimmunity-prone state throughout the lifespan. Although many cytokines elevated in DS have well demonstrated pathogenic roles in the etiology of autoimmune diseases in the general population (e.g., TNF-α, IL6), their consistent upregulation in DS regardless of clinical evidence of autoimmune pathology indicates the existence of a prolonged pre-clinical period, where the hypercytokinemia likely precedes evident tissue damage and symptomology. Alternatively, it is possible that these elevated cytokines are contributing the overall pathophysiology of DS (e.g., cognitive impairments, complications from viral infections) without formal diagnosis of an autoimmune disease. Therefore, measurements of specific immune cell types or cytokines in the bloodstream are unlikely to provide diagnostic value for autoimmunity in DS. However, antigen-specific immune assays, such as T cell or B cell activation assays, may reveal the specific timing of loss of tolerance and transition to clinical phenotypes. Future studies should also include analysis of tissue-resident immune cells, which may identify sites of local autoimmune attack in DS.

Among the many strategies that could be used to attenuate IFN hyperactivity, JAK inhibitors are the most well-studied and have the most approved indications^66^. Of the more than ten globally-approved JAK inhibitors^66^, we chose to employ in our clinical trial the JAK1/3 inhibitor tofacitinib, which is used to treat diverse autoimmune/inflammatory conditions and which was approved in 2020 for treatment of polyarticular course juvenile idiopathic arthritis (pcJIA) in children 2 years and older^66,67^. Notably, all four IFNRs encoded on chr21 utilize JAK1 for signal transduction in combination with either JAK2 or TYK2, making JAK1 inhibitors the most logical choice to dampen the effects of *IFNR* gene triplication. As part of the clinical trial protocol, the approved interim analysis was designed to qualitatively evaluate feasibility and initial safety data on the first 10 participants completing a 16-week course of tofacitinib treatment. This analysis established that there were no AEs that required a change or cessation of tofacitinib dosing and that this medicine is well tolerated in individuals with DS. The clear benefits observed for diverse autoimmune skin conditions align with an increasing body of evidence supporting the use of JAK inhibition for immunodermatological conditions, including their recent approval for alopecia areata and atopic dermatitis in the general population^68,69^. At this sample size, the effects of tofacitinib on HS are inconclusive. Although some participants and caregivers reported benefits in terms of fewer flares of lesser severity, the MSS metric did not show a clear trend, which may reveal the need for more frequent or different types of monitoring for HS, a condition that cycles periodically in severity. Our results indicate that tofacitinib does not fully suppress the immune response in people with DS, but rather attenuates IFN scores and cytokine scores to levels observed in the general population, which is an important consideration given the likely requirement for long-term use of the drug in this population. Furthermore, the effects of the drug are clearly gene-specific, highlighting the presence of inflammatory processes that may not be attenuated with this inhibitor, which could be beneficial in terms of preserving immune activity. Importantly, during treatment, both IFN scores and cytokine scores remain sensitive to immune stimuli, as evidenced by participants who had received a vaccine or experienced an URI before a blood draw (**Figure 6a**, **Figure 6 – figure supplement 1f**). Overall, it is encouraging that key inflammatory markers decreased in a relatively short timeframe, likely offering systemic benefits beyond skin pathology. Importantly, the fact that levels of IL-6 and TNF-α are reduced upon tofacitinib treatment supports the use of JAK inhibitors over TNF-blockers or anti-IL-6 agents in this population. Although TNF-α-blockers are recommended to be used first in the treatment of rheumatoid arthritis in the general population^70^, the value of this recommendation in people with DS remains to be defined. The clear decrease in anti-TPO and anti-TG levels indicates that autoreactive B cell function requires elevated JAK/STAT signaling, but whether this effect is cell-autonomous versus a consequence of a reduced systemic inflammatory milieu will require further investigation. Defining the effect of tofacitinib on other autoantibodies elevated in DS will also require a larger sample size and may be revealed in the full dataset after completion of this trial, along with analysis of potential remodeling of the B cell lineage upon JAK inhibition, such as effects on mature B cells and plasmablast populations.

Lastly, this ongoing clinical trial includes measurements of various dimensions of neurological function not reported here. Although the absence of a placebo control arm may impede a clear interpretation of any effect of JAK inhibition on cognitive function, preliminary results have prompted the design and launch of a second trial (NCT05662228) aimed at defining the relative safety and efficacy of tofacitinib, intravenous immunoglobulin (IVIG), and the benzodiazepine lorazepam for Down syndrome Regression Disorder (DSRD), a condition characterized by sudden loss of neurological function^71^.

Altogether, these findings justify both a deeper investigation of all the deleterious effects of autoimmunity and hyperinflammation in DS and the expanded testing of immunomodulatory strategies for diverse aspects of DS pathophysiology, even perhaps from an early age.

## Materials and Methods

### Human Trisome Project (HTP) study

All aspects of this study were conducted in accordance with the Declaration of Helsinki under protocols approved by the Colorado Multiple Institutional Review Board. Results and analyses presented herein are part of a nested study within the Crnic Institute’s Human Trisome Project (HTP, NCT02864108, see also www.trisome.org) cohort study. All study participants, or their guardian/legally authorized representative, provided written informed consent. The HTP study has generated multiple multi-omics datasets on hundreds of research participants, some of which have been analyzed in previous studies, including whole blood transcriptome data^44^, white blood cell transcriptome data^19^, plasma proteomics^44^, plasma metabolomics^19,44^, and immune mapping via flow cytometry^16^ and mass cytometry^44,50^. This paper reports new analyses of select previous datasets (transcriptome, mass cytometry, MSD immune markers) within the larger multi-omics dataset of the HTP study, as well as analyses of new datasets (e.g., anti-TPO, ANA, autoantibodies), as described in detail below.

### Annotation of co-occurring conditions

Within the HTP, a clinical history for each participant is curated from both medical records and participant/family reports. Both surveys are set up as REDCap^72^ instruments that collect information as a review of systems (e.g., cardiovascular, immunity, endocrine). Expert data curators complete the medical record review and evaluate answers provided by self-advocates and caregivers. In cases of discordant answers across the two instruments, medical records take precedence. De-identified demographic and clinical metadata obtained is then linked to de-identified biospecimens used to generate the various -omics (e.g., RNA sequencing) and targeted assay datasets (e.g., anti-TPO assays). For annotation of AITD, several possible entries were considered as shown in **Figure 1 – figure supplement 1a**, including history of hypothyroidism, hyperthyroidism, Hashimoto’s disease, Grave’s disease, anti-TPO or -TG antibodies, and subclinical hypothyroidism. For annotation of immune skin conditions, atopic dermatitis and eczema were combined and counted in a single group, as were hidradenitis suppurativa (HS), folliculitis, and ‘boils’.

### Blood sample collection and processing

The biological datasets analyzed herein were derived from peripheral blood samples collected using PAXgene RNA Tubes (Qiagen) and BD Vacutainer K2 EDTA tubes (BD). Whole blood from PAXgene collection tubes was processed for RNA sequencing as described below. Two 0.5 mL aliquots of whole blood were withdrawn from each EDTA tube and processed for mass cytometry as described below. The remaining EDTA blood samples were centrifuged at 700 x g for 15 min to separate plasma, buffy coat containing white blood cells (WBC), and red blood cells (RBCs). Samples were then aliquoted, flash frozen and stored at -80°C until subsequent processing and analysis. Centrifugation and storage of samples took place within 2 hours of collection.

### Measurements of autoantibodies

Anti-TPO status was determined from plasma samples using an electrochemiluminescence-based assay ^73^, and carried out by the Autoantibody/HLA Core Facility of the Barbara Davis Center for Childhood Diabetes at the University of Colorado Anschutz Medical Campus. Sample values were calculated as (sample signal – negative control signal) / (positive control signal – negative control signal), with the threshold (upper limit of normal) for TPO positivity based on the 95^th^ percentile of healthy control samples.

Anti-nuclear antigen (ANA) status was determined from plasma samples using a qualitative ELISA kit (MyBioSource, cat. no. 702970) according to manufacturer instructions, with a sample OD450nm / negative control OD450nm ratio ≥ 2.1 evaluated as positive and a ratio < 2.1 evaluated as negative.

Autoantigen profiling of EDTA plasma samples (50 µL each; T21, n = 120; D21, n = 60) was performed by the Affinity Proteomics unit at SciLifeLab (KTH Royal Institute of Technology, Stockholm, Sweden) using peptide arrays. Antigens were selected to cover potential associations to autoimmune diseases and consisted of 380 peptide fragments covering ∼270 unique proteins (1-5 fragments per protein). Fragments were ∼20-163 amino acids long (median 82). All antigens were expressed in E. coli with a hexahistidyl and albumin binding protein tag (His6ABP). Using, COOH-NH2 chemistry, the analyzed antigens, in addition to controls, were immobilized on color-coded magnetic beads (MagPlex, Luminex). Controls consisted of His6ABP, buffer, rabbit anti-human IgG (loading control, Jackson ImmunoResearch), and Epstein-Barr nuclear antigen 1 (EBNA1, Abcam). Research samples and technical controls (commercial plasma; Seralab) were diluted (1:250) in assay buffer, which consisted of 3% BSA, 5% milk, 0.05% Tween-20, and 160 μg/ml His6ABP tag in PBS. Diluted samples and controls were incubated for 1 hour at room temperature then subsequently incubated with the antigen bead array for 2 hour. The reactions were then fixed for 10 minutes using 0.02% paraformaldehyde, then incubated for 30 minutes with goat Fab specific for human IgG Fc-γ tagged with the fluorescent marker R-phycoerythrin (Invitrogen). Median fluorescence intensity (MFI) and number of beads for each reaction was analyzed using a FlexMap 3D instrument (Luminex Corp.). Quality control was performed using MFI and bead count to exclude antigens and samples not passing technical criteria including minimal bead counts and antigen coupling efficiency. To adjust for sample specific backgrounds, MFI values were transformed per reaction median absolute deviations (MADs) using the following calculation:

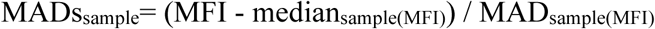

Subsequent data analysis and handling was performed using R. For each antigen, positivity was defined as >90th percentile MAD value for D21 samples only. Overrepresentation of positivity for each antigen in the T21 versus D21 group was determined using Fisher’s exact test, excluding antigens detected in <18 samples (<10% of total experiment). Correction for multiple testing was performed using the Benjamini-Hochberg approach and significance defined as q<0.1 (10% FDR). Similarly, within the T21 group, Fisher’s exact test was used to test for overrepresentation of antigen positivity in cases versus controls for co-occurring conditions, with only those with at least 5 cases considered in the analysis.

### Immune profiling via mass cytometry

Generation of the mass cytometry dataset was described previously^44^, but a full description is included here for reference. Two 0.5 mL aliquots of EDTA whole blood samples underwent RBC lysis and white blood cell fixation using TFP FixPerm Buffer (Transcription Factor Phospho Buffer Set, BD Biosciences). WBCs were then washed in 1x in PBS (Rockland), resuspended in Cell Staining Buffer (Fluidigm) and stored at −80°C. For antibody staining, samples were thawed at room temperature, washed in Cell Staining Buffer, barcoded using a Cell-ID 20-Plex Pd Barcoding Kit (Fluidigm), and combined per batch. Each batch was able to accommodate 19 samples with a common reference sample. Antibodies were either purchased pre-conjugated to metal isotopes or conjugation was performed in-house using a Maxpar Antibody Labeling Kit (Fluidigm). See **Supplementary file 7** for antibodies. Working dilutions for antibody staining were titrated and validated using the common reference sample and comparison to relative frequencies obtained by independent flow cytometry analysis. Surface marker staining was carried out for 30 min at 4°C in Cell Staining Buffer with added Fc Receptor Binding Inhibitor (eBioscience/ThermoFisher Scientific). Staining was followed by a wash in Cell Staining Buffer. Next, cells were permeabilized in Buffer III (Transcription Factor Phospho Buffer Set, BD Pharmingen) for 20 min at 4°C followed by washing with perm/wash buffer (Transcription Factor Phospho Buffer Set, BD Pharmingen). Intracellular transcription factor and phospho-epitope staining was carried out for 1 hour at 4°C in perm/wash buffer (Transcription Factor Phospho Buffer Set, BD Pharmingen), followed by a wash with Cell Staining Buffer. Cell-ID Intercalator-Ir (Fluidigm) was used to label barcoded and stained cells. Labeled cells were analyzed on a Helios instrument (Fluidigm). Mass cytometry data were exported as v3.0 FCS files for pre-processing and analysis.

#### Analysis of mass cytometry data

##### Pre-processing

Bead-based normalization via polystyrene beads embedded with lanthanides, both within and between batches, followed by bead removal was carried out as previously described using the Matlab- based Normalizer tool^74^. Batched FCS files were demultiplexed using the Matlab-based Single Cell Debarcoder tool^75^. Reference-based normalization of individual samples across batches against the common reference sample was then carried out using the R script *BatchAdjust()*. For the analyses described in this manuscript, CellEngine (CellCarta) was used to gate and export per-sample FCS files at four levels: Firstly, CD3+CD19+ doublets were excluded and remaining cells exported as ‘Live’ cells; Live cells were then gated for hematopoietic lineage (CD45-positive) non-granulocytic (CD66-low) cells and exported as CD45+CD66low. Lastly, CD45+CD66low cells were gated on CD3-positivity and CD19- positivity and exported as T- and B-cells, respectively. Per-sample FCS files were then subsampled to a maximum of 50,000 events per file for subsequent analysis.

##### Unsupervised clustering

For each of the four levels (live, non-granulocytes, T cells, and B cells), all 388 per-sample FCS files were imported into R as a flowSet object using the *read.flowSet()* function from the flowCore R package^76^. Next a SingleCellExperiment object was constructed from the flowSet object using the *prepData()* function from the CATALYST package^77^. Arcsinh transformation was applied to marker expression data with cofactor values ranging from ∼0.2 to ∼15 to give optimal separation of positive and negative populations for each marker, using the *estParamFlowVS()* function from the flowVS R package^78^ and based on visual inspection of marker histograms (see **Supplementary file 7)**. Quality control and diagnostic plots were examined with the help of functions from CATALYST and the tidySingleCellExperiment R package. Unsupervised clustering using the FlowSOM algorithm^45^ was carried out using the *cluster()* function from CATALYST, with grid size set to 10 x 10 to give 100 initial clusters and a maxK value of 40 was explored for subsequent meta-clustering using the ConsensusClusterPlus algorithm. Examination of delta area and minimal spanning tree plots indicated that 30-40 meta clusters gave a reasonable compromise between gains in cluster stability and number of clusters for each level. Each clustering level was re-run with multiple random seed values to ensure consistent results.

##### Visualization using t-distributed stochastic neighbor imbedding (tSNE)

Dimensionality reduction to two dimensions was carried out using the *runDR()* function from the CATALYST package, with 500 cells per sample, and using several random seed values to ensure consistent results. Multiple values of the perplexity parameter were tested, with a setting of 440, using the formula Perplexity=N^(1/2) as suggested at https://towardsdatascience.com/how-to-tune-hyperparameters-of-tsne-7c0596a18868, providing a visualization with good agreement with the clusters defined by FlowSOM.

##### Cell type classification

To aid in assignment of clusters to specific lineages and cell types, the MEM package (marker enrichment modeling) was used to call positive and negative markers for each cell cluster based on marker expression distributions across clusters. Manual review and comparison to marker expression histograms, as well as minimal spanning tree plots and tSNE plots colored by marker expression, allowed for high-confidence assignment of most clusters to specific cell types. Clusters that were insufficiently distinguishable were merged into their nearest cluster based on the minimal spanning tree. Relative frequencies for each cell type / cluster were calculated for each sample as a percentage of total live cells and as a percentage of cells used for each level of clustering: total CD45+CD66low cells, total T cells, or total B cells.

##### Beta regression analysis

To identify cell clusters for which relative frequencies are associated with either trisomy 21 status or with various clinical subgroups (e.g. ANA+) among individuals with trisomy 21, beta regression analysis was carried out using the betareg R package, with each model using cell type cluster proportions (relative frequency) as the outcome/dependent variable and either T21 status or clinical subgroups as independent/predictor variables, along with adjustment for age and sex, and a logit link function. Extreme outliers were classified per-karyotype and per-cluster as measurements more than three times the interquartile range below or above the first and third quartiles, respectively (below Q1 - 3*IQR or above Q3 + 3*IQR) and excluded from beta regression analysis. Correction for multiple comparisons was performed using the Benjamini-Hochberg (FDR) approach. Effect sizes (as fold-change in T21 vs. euploid controls or among T21 subgroups) for each cell type cluster were obtained by exponentiation of beta regression model coefficients. Fold-changes were visualized by overlaying on tSNE plots using ggplot2. For visualization of individual clusters, data points were adjusted for age and sex, using the *adjust()* function from the datawizard R package, and visualized as sina plots (separated by T21 status or clinical subgroup).

### Measurement of immune markers and calculation of cytokine scores

Briefly, from each EDTA plasma sample, two replicates of 12-25 µL were analyzed using the Meso Scale Discovery (MSD) multiplex immunoassay platform V-PLEX Human Biomarker 54-Plex Kit (HTP cohort) or U-PLEX Human Biomarker Group 1 71-Plex and V-PLEX Human Vascular Injury Panel 2 Kits (clinical trial cohort) on a MESO QuickPlex SQ 120 instrument. Assays were carried out as per manufacturer instructions. Concentration values were calculated against a standard curve with provided calibrators. MSD data are reported as concentration values in picograms per milliliter of plasma.

#### Analysis of immune marker data

Plasma concentration values (pg/mL) for each of the cytokines and related immune factors measured across multiple MSD assay plates was imported to R, combined, and analytes with >10% of values outside of detection or fit curve range flagged. For each analyte, missing values were replaced with either the minimum (if below fit curve range) or maximum (if above fit curve range) calculated concentration per plate/batch and means of duplicate wells used for subsequent analysis. For the HTP study analysis, extreme outliers were classified per-karyotype and per-analyte as measurements more than three times the interquartile range below or above the first and third quartiles, respectively, and excluded from further analysis. Differential abundance analysis for inflammatory markers measured by MSD was performed using mixed effects linear regression as implemented in the *lmer()* function from the lmerTest R package (v3.1-2) with log2-transformed concentration as the outcome/dependent variable, T21 status or clinical subgroup (e.g., ANA+) as the predictor/independent variable, age and sex as fixed covariates, and sample source as a random effect. Multiple hypothesis correction was performed with the Benjamini-Hochberg method using a false discovery rate (FDR) threshold of 10% (q < 0.1). Prior to visualization or correlation analysis, MSD data were adjusted for age, sex, and sample source using the *removeBatchEffect()* function from the limma package (v3.44.3).

#### Calculation of cytokine scores

For comparison of clinical trial samples across time points, cytokine scores were calculated as the sum of the Z-scores for TNF-α, IL-6, CRP and IP-10. For comparison of clinical trial samples to the HTP cohort, Z-scores were first calculated from age-, sex, and batch-adjusted values for each sample, based on the mean and standard deviation of the HTP euploid control samples.

### Whole blood transcriptome analysis and calculation of IFN scores

Strand-specific sequencing libraries were prepared from globin-depleted, polyA-enriched whole blood RNA and sequenced on the Illumina NovaSeq platform (2x 150 bases). Data quality was assessed using FASTQC (v0.11.5) and FastQ Screen (v0.11.0). Trimming and filtering of low-quality reads was performed using bbduk from BBTools (v37.99) and fastq-mcf from ea-utils (v1.05). Alignment to the human reference genome (GRCh38) was carried out using HISAT2 (v2.1.0) in paired, spliced-alignment mode against a GRCh38 index and Gencode v33 basic annotation GTF, and alignments were sorted and filtered for mapping quality (MAPQ > 10) using Samtools (v1.5). Gene-level count data were quantified using HTSeq-count (v0.6.1) with the following options (--stranded=reverse –minaqual=10 –type=exon -- mode=intersection-nonempty) using a Gencode v33 GTF annotation file. Differential gene expression in T21 versus D21 was evaluated using DESeq2 (version 1.28.1)^79^, with q < 0.1 (10% FDR) as the threshold for differential expression.

#### DS IFN scores

RNA-seq-based ‘Down syndrome interferon scores’ (DS IFN scores) were calculated as follows: for comparison of clinical trial samples across time points, DS IFN scores were calculated as the sum of Z-scores across 16 interferon-stimulated genes (ISGs) genes with significant mean fold-change of at least 1.5 in the HTP T21 group vs. the euploid control group, excluding *IFNAR2*, *MX1*, and *MX2* which are encoded on chromosome 21. For comparison of clinical trial samples to the HTP cohort, gene-wise Z- scores were first calculated from age-, sex, and sequencing batch-adjusted FPKM values for each sample, based on the mean and standard deviation of the HTP euploid control samples.

#### Gene set enrichment analysis (GSEA)

GSEA^80^ was carried out in R using the fgsea package (v1.14.0), using Hallmark gene sets, log2-transformed fold-change values as the ranking metric.

### Clinical trial design and oversight

All aspects of this study were conducted in accordance with the Declaration of Helsinki. All study activities were approved by the Colorado Multiple Institutional Review Board (COMIRB, protocol # 19- 1362, NCT04246372) with an independent Data and Safety Monitoring Board (DSMB) appointed by the National Institute of Arthritis and Musculoskeletal and Skin Diseases (NIAMS). Written consent was obtained from all participants, or their legally authorized representative if the participant was unable to provide consent, in which case participant assent was obtained. The Clinical Trial Protocol is provided in the **Supplementary file 8**. We report here interim results of a single-site, open-label phase 2 clinical trial enrolling individuals with DS between the ages of 12 and 50 years old with moderate-to-severe alopecia areata, hidradenitis suppurativa, psoriasis, atopic dermatitis, or vitiligo. Qualifying disease scores are shown in **Supplementary file 3**. After screening, qualifying participants are prescribed 5 mg tofacitinib twice daily for 16 weeks, with an optional extension arm to week 40. During the main 16-week trial, participants attend five safety monitoring visits after enrollment at the Baseline visit.

### Trial Population

The recruitment goal for this trial is 40 participants completing 16 weeks of tofacitinib treatment, with a qualitative interim analysis triggered when 10 participants completed the main 16-week trial. Of the 10 participants included in the interim analysis, 4 are female, 100% identify as White/Caucasian, 3 identify as Hispanic or Latino, and mean age at enrollment was 23.1 years old (range 15-38.1 years old) (**Supplementary file 3**). Baseline qualifying conditions of the 10 participants were alopecia areata: n=6 (46.1%), hidradenitis suppurativa: n=3 (30.8%), or psoriasis: n=1 (7.7%). Two participants also had atopic dermatitis, and two others had vitiligo, albeit below the severity required to be the qualifying condition. **Outcome Measures.**

#### Primary endpoints

The two primary outcome measures for this trial are safety and reduction in IFN transcriptional scores derived from peripheral whole blood. Based on the safety profile for tofacitinib in the general population^70^, the safety primary endpoint was defined as no more than two serious adverse events (SAEs) definitely attributable to tofacitinib over the course of 16 weeks for 40 participants. Adverse events were classified based using Common Terminology Criteria for Adverse Events 5.0 (CTCAE 5.0). IFN scores are commonly used to monitor disease severity and response to treatment in IFN-driven pathologies^81,82^ and their calculation form RNAseq data is described above.

#### Secondary endpoints

The secondary outcome measures for this trial include improvements in skin health as defined by a global assessment, the Investigator Global Assessment (IGA), as well as the disease- specific assessments. Overall skin pathology, accounting for all present skin conditions regardless of severity, was assessed using a modified IGA which scores on a five-point scale for each skin condition (six points for HS) with a range of 0-21. Another secondary endpoint assessing global skin health is a change in the Dermatological Quality of Life Index (DLQI), used to assess participant-reported impact of skin conditions on self-image, relationships, and daily activities. Possible total scores range from 0-30, with higher scores indicating a more impaired quality of life. Condition-specific assessments used are Severity of Alopecia Tool (SALT) for AA affecting at least 25% of the scalp (qualifying score is ≥ 25); Hidradenitis Suppurativa-Physicians Global Assessment (HS-PGA) to define eligibility (qualifying score ≥3) and Modified Sartorius Scale (MSS) to monitor changes throughout the study for HS; Psoriasis Area and Severity Index (PASI, qualifying score is ≥10) for psoriasis; Vitiligo Extent Tensity Index (VETI, qualifying score is ≥2), for moderate-to-severe vitiligo; and Eczema Area and Severity Index (EASI, qualifying EASI score ≥16) for moderate-to-severe atopic dermatitis. The last secondary endpoint is reduction in a cytokine score coalescing information on four inflammatory markers elevated in DS: Tumor Necrosis Factor Alpha (TNF-α), interleukin 6 (IL-6), C-reactive protein (CRP), and IFN-inducible protein 10 (IP10, CXCL10)^18^. Measurement of these proteins and calculation of the cytokine score is described above.

#### Tertiary endpoints

This clinical trial includes multiple exploratory tertiary endpoints (see full protocol in **Supplementary file 8**), including reduction in autoantibodies related to AITD (anti-TPO, anti-TG, and anti-TSHr) and celiac disease (anti-tTG, anti-DGP). In the clinical trial, these autoantibodies were assessed using established clinical assays.

### Statistical Analysis

The Statistical Analysis Plan (SAP) approved by the appointed DSMB is included with the Clinical Trial Protocol in the **Supplementary file 8**. This report includes analysis of the time points used to assess endpoints (baseline and 16 weeks), as well research-only time points at 2 and 8 weeks of treatment. Given the qualitative nature of this interim analysis, statistical analysis is not completed for changes observed between baseline and the 16-week endpoint. Data may be displayed as log2 transformed for clarity in viewing the graphs.

## Data Availability Statement

A collection with datasets used in this study is available in Synapse (https://doi.org/10.7303/syn53185135), with the individual datasets also accessible as detailed below.

Demographic and health history data for research participants in the HTP study are available on both the Synapse data sharing platform (https://doi.org/10.7303/syn31488784) and through the INCLUDE Data Hub (https://portal.includedcc.org/). Mass cytometry data for 380+ HTP research participants are available both in Synapse (https://doi.org/10.7303/syn53185253) and FlowRepository, Study ID FR-FCM-Z5GE, https://flowrepository.org/id/RvFrQaBqhe8TGyko1OMdQKtR7HN8nulAnHW0PJkm1CEyyo8fnJg2rHrWvNrhE8xu. Targeted plasma proteomics for inflammatory markers using Meso Scale Discovery (MSD) assays for 470+ HTP research participants can be accessed through Synapse (https://doi.org/10.7303/syn31475487) and the INCLUDE Data Hub. Whole blood transcriptome data for 400 HTP research participants can be accessed through Synapse (https://doi.org/10.7303/syn31488780), the INCLUDE Data Hub, and NCBI Gene Expression Omnibus (GSE190125). Whole blood transcriptome data for 10 clinical trial participants at baseline and after 2, 8, and 16 weeks of tofacitinib treatment can be accessed through Synapse (https://doi.org/10.7303/syn53185250), and NCBI Gene Expression Omnibus (GSE PENDING). Targeted plasma proteomics for inflammatory markers using Meso Scale Discovery (MSD) assays for 10 clinical trial participants can be accessed through Synapse (https://doi.org/10.7303/syn53185252).

## Code Availability Statement

No custom code or algorithms were developed during the course of this study. R analysis scripts will be made available upon request.

## Acknowledgements

This work was supported primarily by NIH grant R61AR077495. Additional funding was provided by NIH grants R01AI150305 (J.M.E.), T32CA190216 (K.A.W.), 2T32AR007411- 31 (K.A.W.), UM1TR004399 (data generation and REDCap support), P30CA046934 (support of shared resources), the Linda Crnic Institute for Down Syndrome, the Global Down Syndrome Foundation, the Anna and John J. Sie Foundation, the Human Immunology and Immunotherapy Initiative, the University of Colorado School of Medicine, the Boettcher Foundation, and Fast Grants. We are grateful to all research participants and their families involved in the Human Trisome Project and the clinical trial. We thank Lyndy Bush for administrative support, Dr. Kim Jordan and her team at the Human Immune Monitoring Shared Resource for outstanding service in generation of the immune marker dataset, and Dr. Eric Clambey and his team at the Flow Cytometry Shared Resource for outstanding service in generation of the mass cytometry dataset. We are also grateful to the Colorado Translational and Sciences Institute and the Colorado Multiple Institutional Review Board for assistance in all clinical research projects involving the Crnic Institute. Special thanks to Michelle Sie Whitten, the team at the Global Down Syndrome Foundation, Dr. John Reilly, and Dr. Ron Sokol for logistical support at multiple stages of the project.

## Author Contributions Statement

A.L.R.: conceptualization, methodology, formal analysis, project administration, data curation, visualization, investigation, resources, writing of manuscript; E.W.: methodology, data curation, investigation, writing of manuscript; E.G.: methodology, data curation, investigation, writing of manuscript; BEE: methodology, project administration, data curation, investigation, writing of manuscript; K.R.W.: methodology, data curation, investigation, writing of manuscript; K.P.S.: methodology, project administration, formal analysis, data curation, visualization, investigation, writing of manuscript; P.A.: conceptualization, methodology, formal analysis, data curation, visualization, investigation, writing of manuscript; K.A.W.: conceptualization, methodology, investigation, writing of manuscript; R.E.G.: methodology, investigation, writing of manuscript; E.B.: methodology, investigation, writing of manuscript; H.R.L.: methodology, investigation, writing of manuscript; M.G.D.: methodology, investigation, formal analysis, data curation, visualization, writing of manuscript; N.P.E.: methodology, investigation, formal analysis, data curation, visualization, writing of manuscript; A.A.H.: conceptualization, methodology, project administration, investigation, writing of manuscript; B.M.: methodology, data curation, investigation, writing of manuscript; K.D.S.: methodology, investigation, visualization, writing of manuscript; L.P.: methodology, investigation, formal analysis, data curation, project administration, writing of manuscript; D.F.: methodology, investigation, formal analysis, data curation, project administration, writing of manuscript; M.D.G.: methodology, investigation, formal analysis, data curation, visualization, project administration, writing of manuscript; C.A.D.: conceptualization, methodology, data curation, investigation, project administration, writing of manuscript; D.A.N.: conceptualization, methodology, data curation, investigation, resources, project administration, writing of manuscript; J.M.E.: conceptualization, methodology, project administration, visualization, investigation, resources, writing of manuscript.

## Supplementary File Legends

**Supplementary file 1.** (A) Cohort characteristics and (B) clinical data for Human Trisome Project participants analyzed in this study.

**Supplementary file 2.** Autoantibody measurements of Human Trisome Project participants: (A) anti- thyroid peroxidase (TPO) reactivity; (B) anti-nuclear antigen (ANA) reactivity; (C) SciLifeLabs autoantigen peptide array data.

**Supplementary file 3.** (A) Minimum qualifying scores for skin conditions. (B) Cohort characteristics for clinical trial participants.

**Supplementary file 4.** Adverse events for clinical trial participants.

**Supplementary file 5.** Skin pathology metrics for clinical trial participants: (A) Investigator’s Global Assessment (IGA); (B) Dermatology Life Quality Index (DLQI); (C) Severity of Alopecia Tool (SALT); (D) Psoriasis Area and Severity Index (PASI); and (E) Eczema Area and Severity Index (EASI).

**Supplementary file 6.** (A) DS IFN scores; (B) Cytokine scores; (C) anti-thyroid peroxidase (TPO) titers; (D) anti-transglutaminase (TG) titers; and (E) anti-thyroid stimulating hormone receptor (TSHR) titers for clinical trial participants.

**Supplementary file 7.** Marker information for mass cytometry analysis.

**Supplementary file 8.** Clinical trial protocol.

